# De novo variants in *KDM2A* cause a syndromic neurodevelopmental disorder

**DOI:** 10.1101/2025.03.31.25324695

**Authors:** Eric N. Anderson, Stephan Drukewitz, Sukhleen Kour, Anuradha V. Chimata, Deepa S. Rajan, Senta Schönnagel, Karen L. Stals, Deirdre Donnelly, Siobhan O’Sullivan, John F. Mantovani, Tiong Y. Tan, Zornitza Stark, Pia Zacher, Nicolas Chatron, Pauline Monin, Severine Drunat, Yoann Vial, Xenia Latypova, Jonathan Levy, Alain Verloes, Jennefer N. Carter, Devon E. Bonner, Suma P. Shankar, Jonathan A. Bernstein, Julie S. Cohen, Anne Comi, Deanna Alexis Carere, Lisa M Dyer, Sureni V Mullegama, Pedro A. Sanchez-Lara, Katheryn Grand, Hyung-Goo Kim, Afif Ben-Mahmoud, Sidney M. Gospe, Rebecca S. Belles, Gary Bellus, Klaske D. Lichtenbelt, Renske Oegema, Anita Rauch, Ivan Ivanovski, Frederic Tran Mau-Them, Aurore Garde, Rachel Rabin, John Pappas, Annette E. Bley, Janna Bredow, Timo Wagner, Eva Decker, Carsten Bergmann, Louis Domenach, Henri Margot, Undiagnosed Diseases Network, Johannes R. Lemke, Rami Abou Jamra, Julia Hentschel, Heather Mefford, Amit Singh, Udai Bhan Pandey, Konrad Platzer

## Abstract

Germline variants that disrupt components of the epigenetic machinery cause syndromic neurodevelopmental disorders. Using exome and genome sequencing, we identified *de novo* variants in *KDM2A*, a lysine demethylase crucial for embryonic development, in 18 individuals with developmental delays and/or intellectual disabilities. The severity ranged from learning disabilities to severe intellectual disability. Other core symptoms included feeding difficulties, growth issues such as intrauterine growth restriction, short stature and microcephaly as well as recurrent facial features like epicanthic folds, upslanted palpebral fissures, thin lips, and low-set ears. Expression of human disease-causing *KDM2A* variants in a Drosophila melanogaster model led to neural degeneration, motor defects, and reduced lifespan. Interestingly, pathogenic variants in KDM2A affected physiological attributes including subcellular distribution, expression and stability in human cells. Genetic epistasis experiments indicated that KDM2A variants likely exert their effects through a potential gain-of-function mechanism, as eliminating endogenous KDM2A in Drosophila did not produce noticeable neurodevelopmental phenotypes. Data from Enzymatic-Methylation sequencing supports the suggested gene-disease association by showing an aberrant methylome profiles in affected individuals’ peripheral blood. Combining our genetic, phenotypic and functional findings, we establish *de novo* variants in *KDM2A* as causative for a syndromic neurodevelopmental disorder.

## Introduction

The epigenetic machinery encompasses proteins that function as writers, erasers, readers and remodelers of epigenetic marks on DNA and histones. The eraser KDM2A removes mono- and di-methylation from Histone 3 at Lysine 36 (H3K36). The post-translational demethlyation of lysine residues on histone tails mediated by histone lysine demethylases (KDMs), and specifically by KDM2A, are important players in gene regulation and have shown to be crucial for embryonic development and processes like proliferation, apoptosis and differentiation.^1^

Pathogenic germline variants in genes of the epigenetic machinery cause a now established group of rare Mendelian disorders.^2,3^ Within this broad group, variants disrupting KDMs are a frequent cause of neurodevelopmental disorders (NDDs),^4–6^ which encompass a heterogeneous group of conditions characterized by aberrant brain development and function, leading to cognitive, motor, and behavioral impairments. Previously, two *de novo* missense and one *de novo* frameshift variant in *KDM2A* were described in three individuals with autism and a neurodevelopmental disorder, but only as members of much broader cohorts looking into the genetic architecture of these phenotypes.^7–9^

Here, we describe the overlapping phenotype of 18 individuals with *de novo* variants in *KDM2A*. We combined the power of genetically modified fruit fly, which our lab has successfully used to in the delineation of other novel NDDs^10–12^ with human cell culture, as well as methylome data based on blood-derived DNA from affected individuals, to establish the gene-disease association of *de novo* variants in *KDM2A* and a syndromic neurodevelopmental disorder.

## Subjects and methods

### Recruitment of affected individuals and consent

This study was approved by the ethics committee of the University of Leipzig (402/16-ek). Written informed consent for molecular genetic testing and data publication was obtained from all individuals and/or their legal representatives by the referring physicians according to the guidelines of the ethics committees and institutional review boards of the respective institutes. The compilation of the cohort was supported by international collaboration and online matchmaking via GeneMatcher.^13^ Phenotypic and genotypic information were obtained from the referring collaborators using a standardized questionnaire.

### Variant identification

Trio exome or genome sequencing was performed for all affected individuals and their parents except for individual 12 (duo exome with targeted variant testing of the other parent) and 16 (singleton exome). All individuals were evaluated in the context of local diagnostic protocols. Since no causative variants were identified in a known rare disease gene, research evaluation of the sequencing data was done to potentially identify causative variants in candidate genes. The gnomAD v4 dataset served as the control population.^14^ There were no significant findings, apart from the described variants in *KDM2A*, which likely explain the neurodevelopmental phenotypes of the respective individuals. All variants described were aligned to hg38, mapped to the *KDM2A* MANE Select transcript NM_012308.3 and classified according to ACMG criteria (Table S1).^15,16^

### *In silico* prediction

*M*issense variants were assessed using CADD-v1.6^17^, REVEL^18^ MutPred2,^19^ VEST4^20^ and BayesDel^21^ using deleterious predictions cutoffs defined by Pejaver *et al.*^22^ (Table S2).

### Fly Stock

The KDM2A-WT, KDM2A-P235L, KDM2A-Y141C, and KDM2A-H811N lines were generated by site-specific insertion of the transgene at BestGene Inc using the attP2 insertion vector as previously done.^23^ All Drosophila stocks were maintained on standard cornmeal medium at 29°C in light/dark-controlled incubators. The ELAV-gal4 (#8760), GMR-gal4 (#1104), and Luciferase (#35788) were obtained from the Bloomington Drosophila stock center.

### Climbing assay

The rapid iterative negative geotaxis (RING) assay was done as previously described.^11,24,25^ Briefly, flies expressing KDM2A variants or luciferase pan-neuronally were aged for 20 days before being transferred to fresh vials. Flies were knocked three times on the base of a bench and a video camera was used to record the flies climbing up the wall of the vials. The velocity (cm/s) was calculated and analyzed from three independent experiments using GraphPad Prism 6.

### Lifespan assay

The lifespan assay was performed as previously described.^11,24^ Briefly, flies expressing KDM2A variants or luciferase pan-neuronally were raised on standard cornmeal food. 1–3-day old adult progeny flies were transferred to fresh food twice a week, the number of dead flies were counted every day, and survival functions were calculated and plotted as Kaplan-Meier survival curves. Log-rank with Grehan-Breslow-Wilcoxon tests were performed to determine significance of differences in survival data between the groups using GraphPad Prism 6 software.

### Eye severity experiments in Drosophila

Glass multiple reporter promoter element (GMR-gal4) was used to cross *KDM2A* or luciferase in the eyes. Images of the right eyes from F1 generation adult female Drosophila were taken at day 1 using a Leica M205C dissection microscope equipped with a Leica DFC450 camera. External eye severity was quantified using a previously published scoring system.^26^ Statistical analyses were performed using GraphPad Prism 6 with group comparisons were performed using One-way ANOVA.

### Western blotting

#### Drosophila

On day 1, heads from adulte female F1 generation were collected from each cross and snap-frozen on dry ice. Five heads were used per lane of the western blots. Heads were crushed on dry ice and incubated in RIPA buffer containing 150 mM NaCl, 1% NP40, 0.1% SDS, 1% sodium deoxycholate, 50 mM NaF, 2 mM EDTA, 1 mM DTT, 0.2 mM Na orthovanadate, 1 × protease inhibitor cocktail (Roche; 11836170001). Lysates were sonicated and centrifuged to remove exoskeletal debris. Supernatants were boiled in Laemmli Buffer (Boston Bioproducts; BP-111R) for 5 min and proteins were separated using 3–8%, NuPAGE tris-acetate gels (ThermoFisher Scientific: EA03785BOX). Proteins were transferred onto nitrocellulose membranes (iBlot 2 transfer stacks; Invitrogen, IB23001) using the iBlot2 system (Life Technologies; 13120134). Membranes were blocked in milk (BLOT–QuickBlocker reagent; EMD Millipore; WB57-175GM) and incubated overnight in primary antibody (Rabbit anti-KDM2A, Abcam; ab191387; 1:1000). Blots were washed and incubated in secondary antibody for 1 hour (anti-rabbit, DYLight 800, Pierce, 1:10 000). Imaging was performed using the Odyssey CLx (LI-COR Biosciences). Protein levels were quantified using Image Studio (LI-COR Biosciences), and statistical analyses were performed with GraphPad Prism 6. All Western blots were performed in triplicate using biological replicates.

#### Mammalian cells

Human embryonic kidney 293T (HEK293T) cells from ATCC were cultured in advanced Dulbecco’s Modified Eagle Medium (DMEM) (Gibco; 12491023) containing 10% FBS (Biowest; S01520) and 1 × GlutaMAX (Gibco; 35050079). Cells were lysed by boiling for 5 min in 1 × LDS Sample buffer (Invitrogen; NP-0007) and RIPA buffer. All NuPAGE and western blotting steps were performed as described above. Experiments were performed in triplicate using three independent lysate preparations from cultured cells.

#### Plasmids

KDM2A-WT-HA (VB: 220629-1403rdb), KDM2A-P235L-HA (VB220629-1413ecm), KDM2A-Y141C-HA (VB: 220629-1409axj), and KDM2A-811N-HA (VB:220629-1405kmm) was constructed by VectorBuilder Inc.

#### Immunofluorescence

HEK293T cells transfected with KDM2A plasmid grown on coverslips were rinsed in PBS (Lonza 17-512F) and fixed in 4% paraformaldehyde (Sigma P6148) for 20 min at room temperature. Following fixation, the samples were washed four times (× 10 min) in PBS and blocked with blocking buffer: 5% normal goat serum (NGS; Abcam AB7681) in PBS with 0.1% TritonX-100 (PBST). The samples were incubated overnight at 4 °C with primary antibody Rabbit anti-KDM2A (1:1000) and mouse anti-HA (H3663; 1:1000; Millipore Sigma), washed four times (× 10 min) with 0.1% PBST, and incubated with secondary antibody (goat anti-mouse Alexa Fluor 568; A22287: 1:500 and goat anti-rabbit Alexa Fluor 488; A-11008: 1:500, Invitrogen) for 2 h at room temperature followed by 0.1% PBST washes. Samples were mounted onto slides using Fluoroshield (Sigma F6057).

#### Nuclear-Cytoplasmic Fractionation

HEK293 T cells transfected with KDM2AWT, or KDM2A variants (P235L, Y141C, and H811N) were harvested and nuclear-cytoplasmic fractionation was done using the NE-PER nuclear-cytoplasmic extraction kit per manufacturer’s protocol (ThermoFisher Scientific).

#### Cycloheximide Chase Assay

To evaluate the stability of the KDM2A WT and the P235L variant, HEK293-T cells were transfected with the KDM2A plasmids and incubated for 24 hours. Protein synthesis was then inhibited by addition of 0.5 mg/mL cycloheximide. At 5 different time points (0, 6, 12, 24, and 48 hours after cycloheximide addition) the cells were harvested, and samples’ supernatants were subjected to immunoblot analysis with HA antibody to recognize KDM2A and loading control Tubulin.

### Methylome analysis and Episignature

DNA was extracted from peripheral blood using standard protocols at the respective centers. DNA quantity was measured using Qubit 4 and Qubit dsDNA BR Assay-Kit (ThermoFisher Scientific, Darmstadt, Germany). A total amount of 200 ng DNA was used for ultrasonic fragmentation of the DNA with a focused ultrasonicator (ME220, Covaris®, Massachusetts, United States). Library preparation was done using NEB Enzymatic Methyl-seq Library Preparation Kit. 187.5 ng library was used for capture with the Twist Human Methylome Panel according to the manufactures protocol (TWIST Bioscience, South San Francisco, United States). This panel targets 5.549 million CpG sites (in comparison: on the EPIC v2.0 array 0.930 million CpG sites are covered; 0.163 million CpGs are unique on the array while 4.782 million are unique in the methylome panel). 2×150 bp sequencing was done on one lane of a S4 cartridge (s4 reagent kit (300 cycles), running on a NovaSeq6000 (Illumina, San Diego, CA, United States). The average coverage of targets regions was 120x. As control group, DNA from peripheral blood of healthy age- and sex-matched individuals were selected. Raw reads were quality checked using FastQC and trimmed using cutadapt.^27^ Trimmed data was aligned to GRCh38 using BWA-meth, duplicates were marked using Picard – MarkDuplicates. Methylation calls were extracted from the deduplicated alignment files using MethylDackel. Bedgraphs were filtered for regions covered > 30X. Calling of differential methylated regions (DMR) between case and control group was done using metilene with the following settings, min. length of CpG >=10, min. length in nt > 0, min. absolute methylation difference >=0.1, adjusted p-value <0.05.^28^ To generate an episignature of the *KDM2A* cases, called DMRs were filtered using an adjusted p-value < 0.01 and a minimal absolute methylation difference of 10%.

To visualize the episignature, methylation rates of the DMRs were extracted from the MethylDackel output. Hierarchical clustering and heatmap visualization were performed using the clustermap function implemented from the Seaborn Python package.^29^

## Results

### Clinical description

Here we describe a cohort of 18 individuals including one prenatal case with *de novo* variants in *KDM2A*. An overview of the clinical data on all individuals is presented in Figure 1A and Table 1 (for a detailed phenotypic description see the supplemental case reports and Table S3).

**Figure 1.**
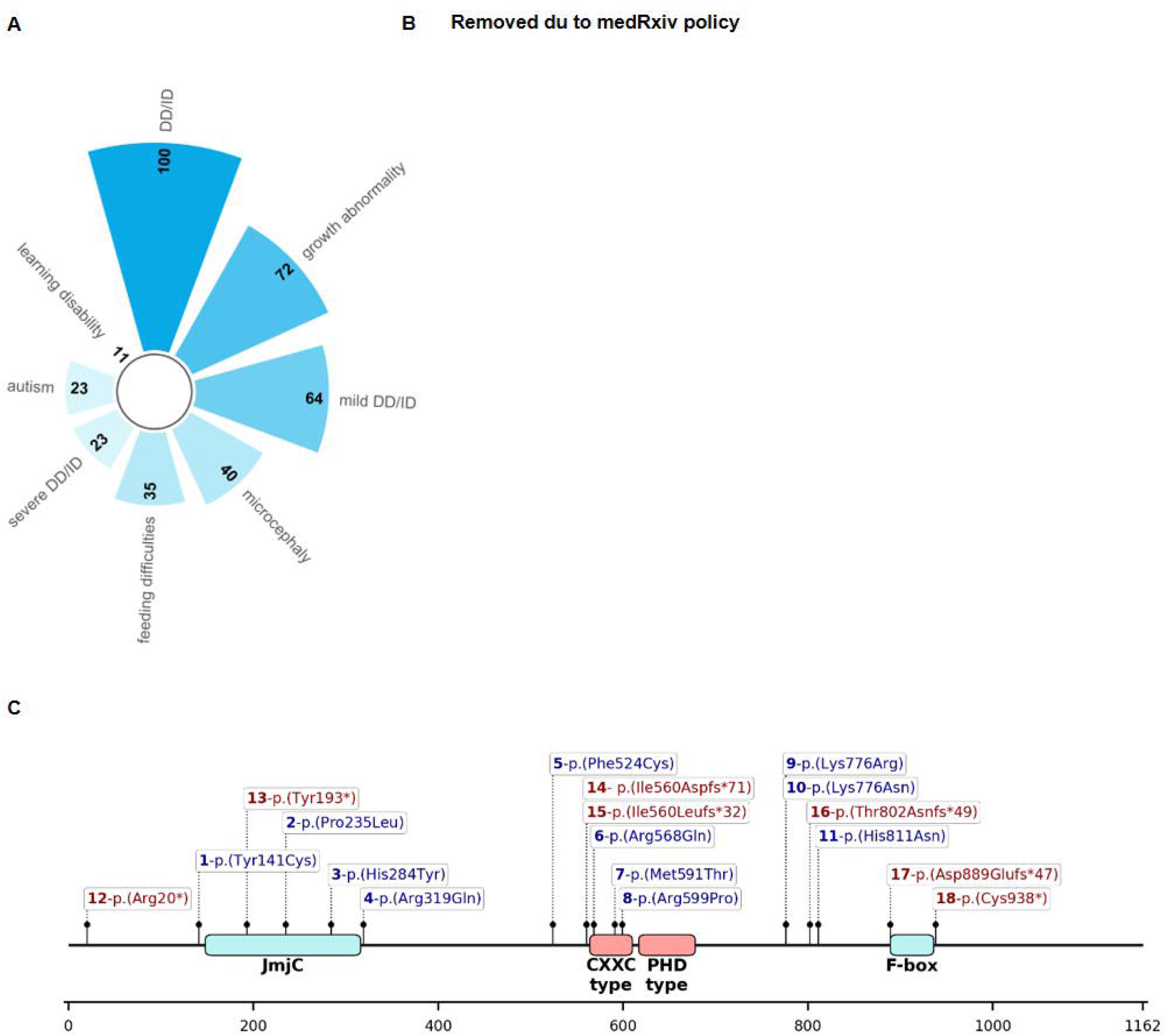
Prevalence of clinical findings and variant location on KDM2A protein level. (A) Radial bar chart illustrating the core symptoms of the *KDM2A*-related neurodevelopmental disorder sorted by frequency. Numbers denote the frequency if each symptom in the cohort. DD: developmental delay; ID: intellectual disability. (B) Facial appearance of individuals at different ages that harbor missense variants or predicted loss-of-function variants in *KDM2A*. Epicanthus, upslanted palpebral fissures, thin upper and/or lower lips and low-set ears were noted as recurrent dysmorphic facial features. (C) Linear schematic representation of the KDM2A protein and location of the variants [GenBank: NM_012308.3]). Bold numbers indicate individual within the cohort. Blue variants represent missense, red variants indicate predicted loss-of-function variants.

**Table 1.**
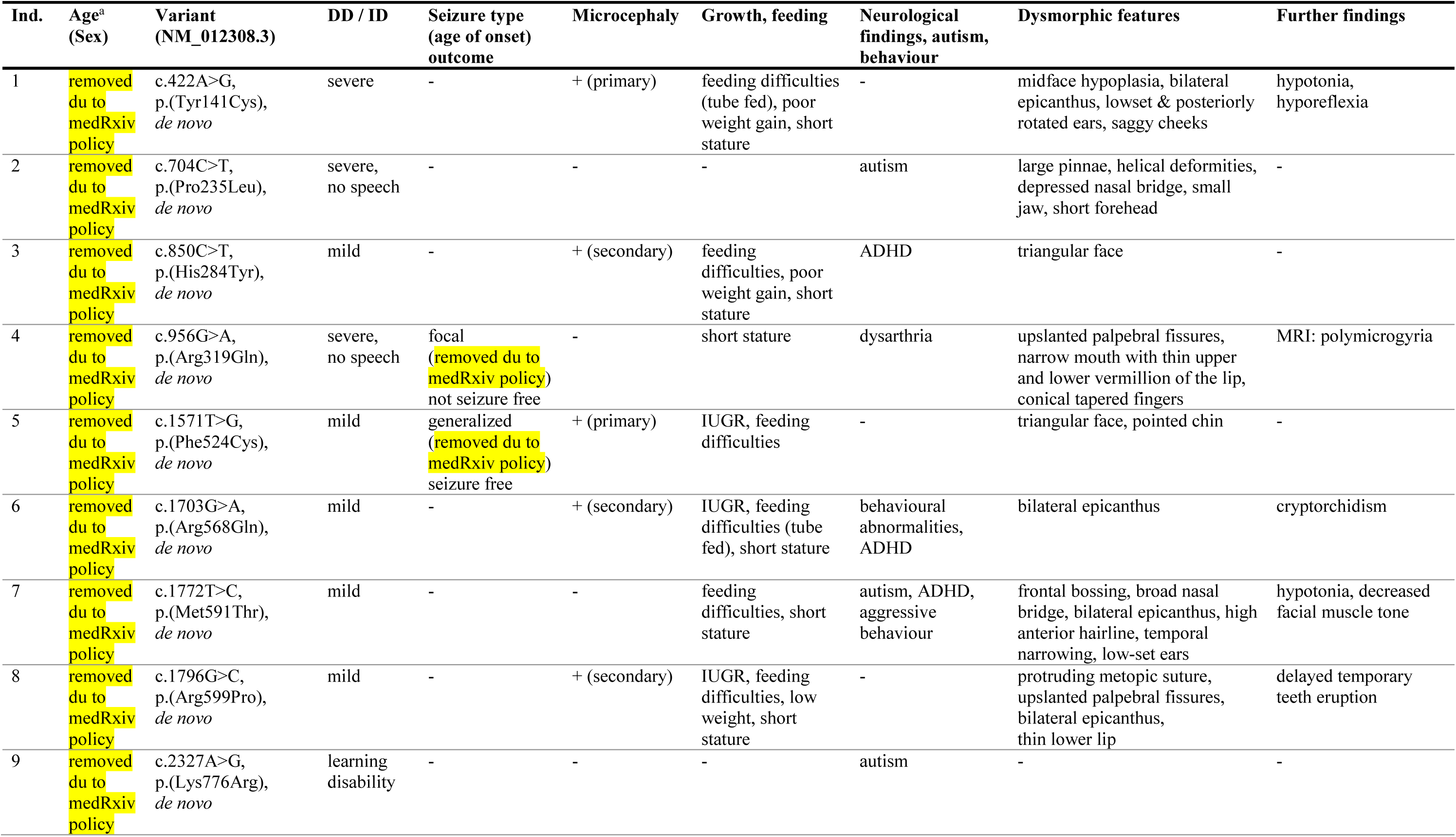
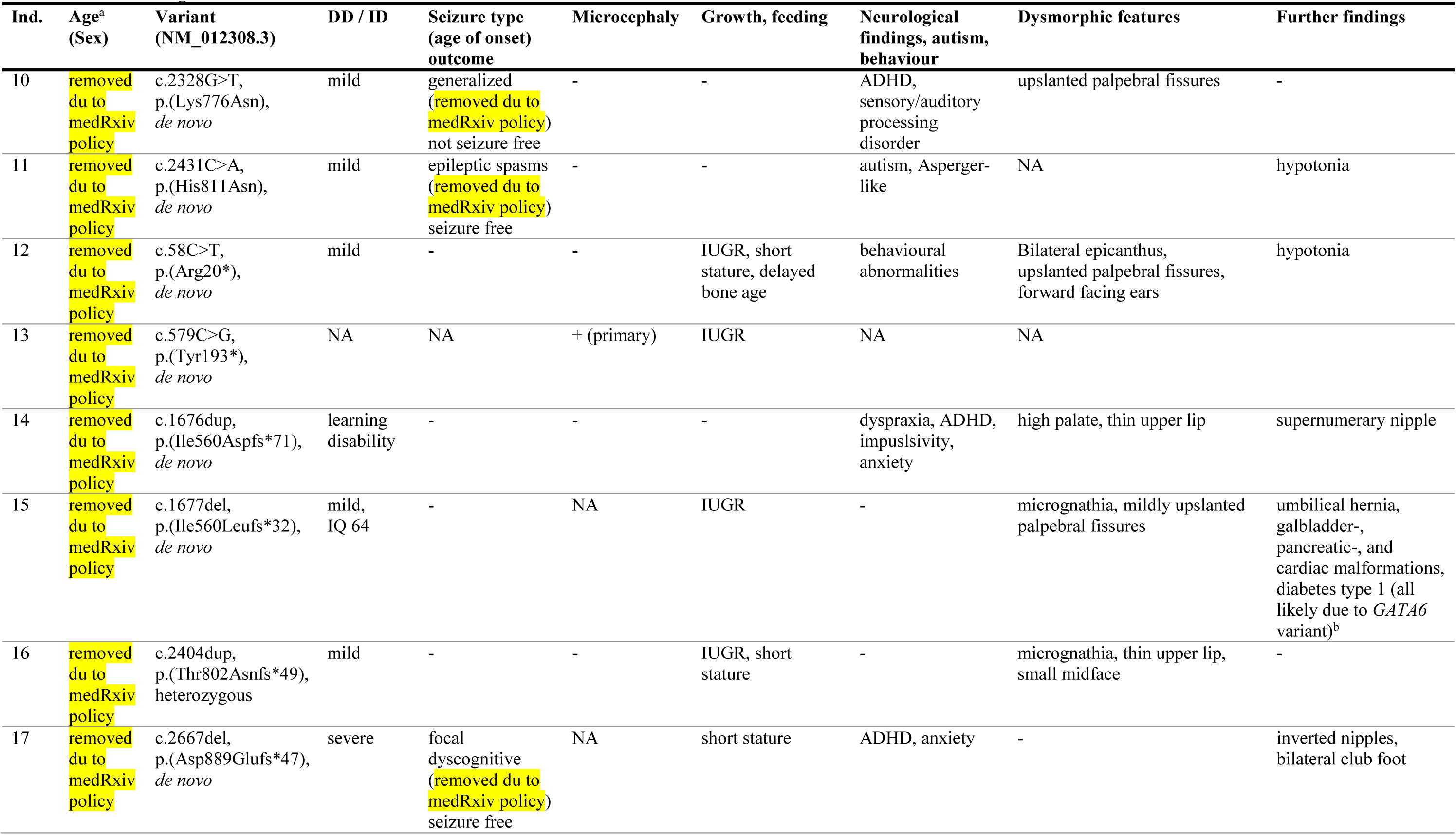
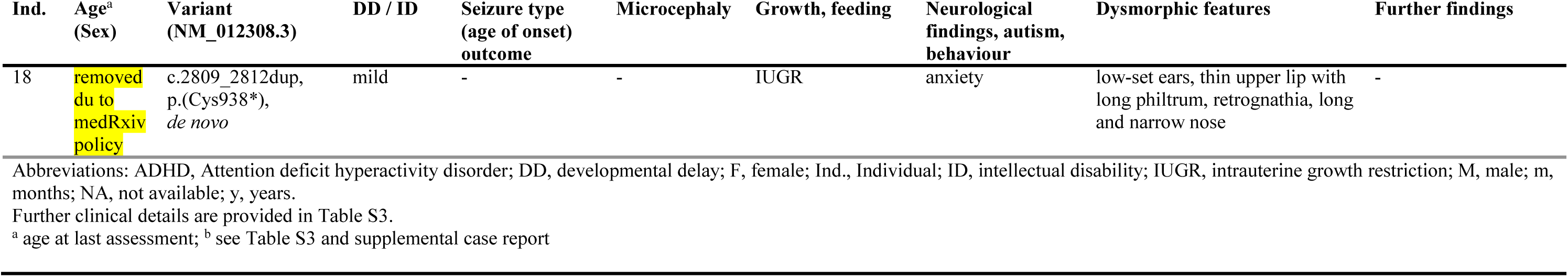
Clinical and genetic details of all affected individuals with causative variants in *KDM2A*.

All individuals, excluding the prenatal case, exhibited developmental delay (DD) and/or intellectual disability (ID) with severity of DD/ID ranging from learning disabilities to severe ID. The majority of individuals were affected by mild DD/ID (11/17) or learning disabilities (2/17), while four individuals presented with severe DD/ID. Microcephaly was observed in six individuals, including the prenatal case. Three of the six individuals presented with primary microcephaly and three with secondary microcephaly. Autism was diagnosed in four individuals, while other behavioural abnormalities such as attention deficit hyperactivity disorder or aggressive behaviour were noted in seven individuals. Seizures ocurred in five individuals with onsets ranging from four months to ten years. Seizure types included focal- and generalized seizures as well as epileptic spasms. At the last assessment, two individuals continued to experience seizures, while three achieved seizure freedom. Hypotonia was reported in four individuals. Cranial MRI was performed in 13 individuals, revealing normal results in seven. Minor and nonspecific abnormalities, such as cerebellar tonsillar ectopia and multiple white matter hyperintensities, were reported in four individuals. Of note, individual 4 did present with a malformation of cortical development of the polymicrogyria spectrum. None of the individuals was reported to show signs of developmental regression. Another notable recurring phenotype in the cohort were growth abnormalities including intrauterine growth restriction (IUGR) and/or short stature in thirteen individuals, including the prenatal case. Eight individuals showed IUGR and nine individuals developed short stature. Of note, four of these thirteen individuals presented with both IUGR and short stature later in life. Feeding difficulties were observed in six individuals, four of whom experienced these issues during the neonatal period, with two requiring tube feeding. Additionally, two individuals had persistent difficulties gaining weight later in life. The referring clinicians reported dysmorphic facial features in twelve individuals: epicanthus, upslanted palpebral fissures, thin upper and/or lower lips and low-set ears emerged as recurrent descriptive features that each occurred in at least three individuals (Figure 1B, Table 1).

### Genetic results

Exome and/or genome sequencing revealed *de novo* variants in *KDM2A* in 17 of the 18 affected individuals (Figure 1C). Individual 16 was identified using a singleton exome, but segregation analysis of the *KDM2A* variant in the parents was not available. We identified eleven distinct *de novo* missense variants and seven predicted loss-of-function (pLoF) variants. None of the variants are recurrent and only two missense variants affect the same amino acid residue p.Lys776. All of the identified variants are absent from gnomAD (v4 dataset).^14^ The eleven missense variants all affect moderate to highly conserved amino acid residues of KDM2A, but *in silico* prediction of potential deleteriousness varies within the cohort. Seven of the missense variants are predicted to be damaging by multiple algorithms, while results of the other four variants encompass mixed and benign predictions (Table S2 and S5).

According to gnomAD, *KDM2A* is a gene with a significantly reduced number of pLoF as well as missense variants, indicating that there is a selective constraint on both types of variants in the general population that lacks severe, early-onset phenotypes such as DD and ID (LOEUF = 0.06; pLI = 1; o/e for missense variants = 0.49; z-score = 6.9).^14^

### P235L variant alters nuclear localisation and decreases KDM2A protein levels

KDM2A protein is primarily localized in the nucleus, and its interaction with unmethylated CpG DNA is crucial for maintaining heterochromosomal homeostasis.^30,31^ To investigate the impact of missense variants on the intracellular localization of KDM2A proteins, we introduced HA-tagged wild-type (WT) and mutant forms (P235L, Y141C, and H811N) of KDM2A into HEK 293T cells. While the expression of WT, Y141C, or H811N did not visibly affect the distribution and localization of KDM2A, the expression of the P235L variant led to the relocation of predominantly nuclear KDM2A to the cytoplasm (Figure 2A, 2B). Remarkably, the expression of the exogenous P235L variant also caused the cytoplasmic mislocalization of endogenous KDM2A (Figure 2A, 2C). To further illustrate the disruption in the localization of KDM2A variants, nucleocytoplasmic fractionation was performed in HEK293T cells expressing exogenous WT or mutant KDM2A proteins, and both exogenous and endogenous KDM2A were probed using Western blotting (Figure 2D). Interestingly, we observed a significant decrease in the nuclear/cytoplasmic ratio in both exogenous and endogenous KDM2A proteins in the P235L variant. In contrast, no such alterations were detected in cells expressing Y141C or WT (Figure 2E, 2F), indicating that the P235L variant interferes with transport of KDM2A into the nucleus.

**Figure 2.**
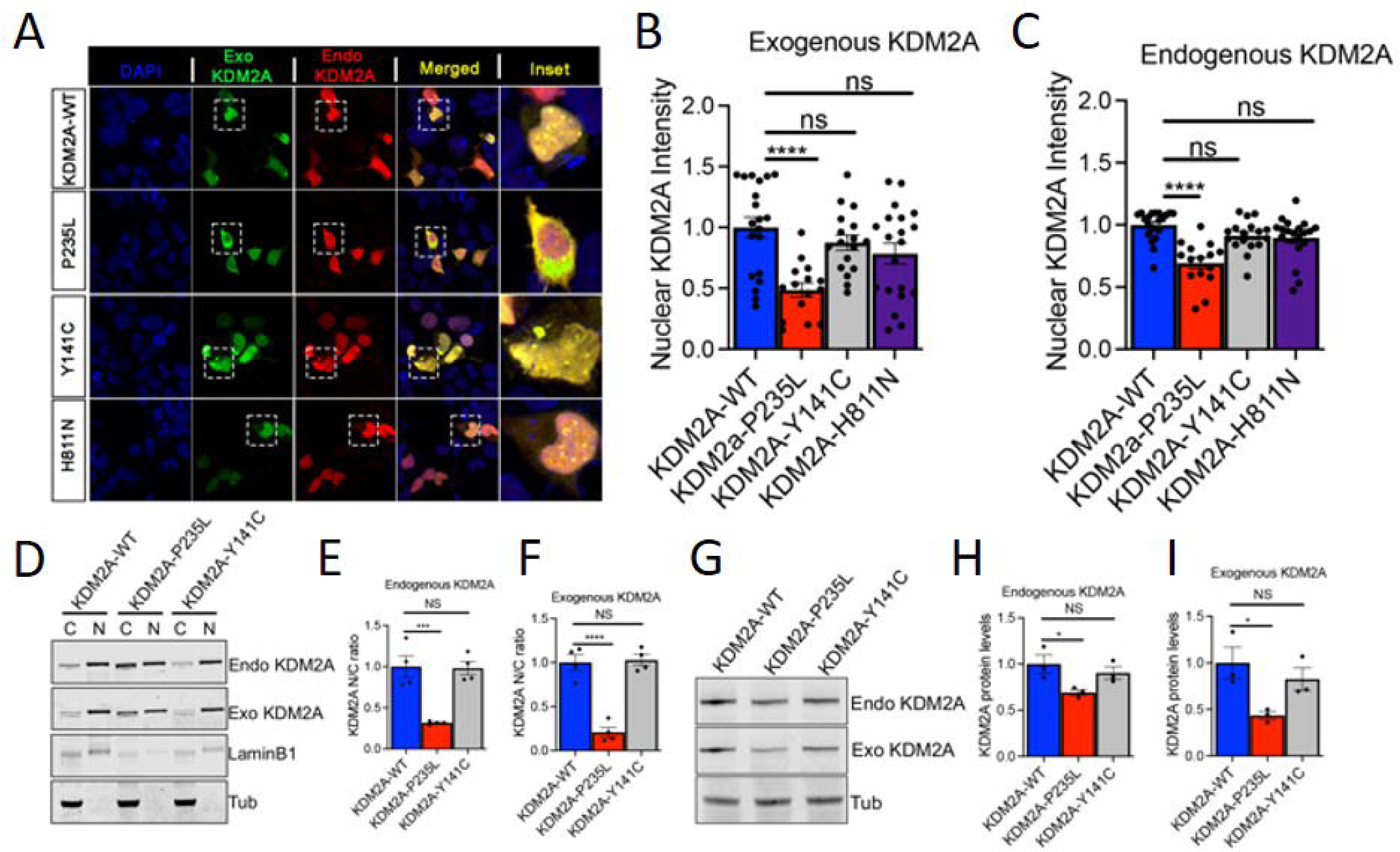
KDM2A variants alter the subcellular distribution of KDM2A protein in mammalian cells. (a) Representative immunofluorescence images of human embryonic kidney cells 293T (HEK293T) transfected with HA-tagged wild type KDM2A (KDM2A-WT) or variants (P235L, Y141C, or H811N) stained for exogenous (green) and endogenous (red) KDM2A protein. DAPi was used to label nuclei. (b) Exogenous KDM2A nuclear intensity quantification showed that the P235L (****p<0.0001) but not the Y141C or H811N showed significantly decreased nuclear intensity compared to exogenous KDM2A-WT (N = 14-20 cells). (c) Endogenous KDM2A nuclear intensity quantification showed that the P235L (****p<0.0001) but not the Y141C or H811N produced a significant reduction in nuclear intensity of endogenous KDM2A protein as compared to KDM2A-WT (N = 14-20 cells). (d) Western blots (WB) of cytoplasmic (C) and nuclear (N) fractions from HEK cells transfected with KDM2A-WT, P235L, and Y141C variants probed for exogenous KDM2A (anti-HA), endogenous KDM2A (anti-KDM2A), nuclear membrane marker (laminB1), and tubulin. (e) Nuclear-cytoplasmic (N/C) ratio quantification of endogenous KDM2A (n = 3 blots, ***p < 0.001). (f) Nuclear-cytoplasmic (N/C) ratio quantification of exogenous KDM2A (N = 3 blots, ***p < 0.001). (g) WB of HEK cells transfected with KDM2A-WT, P235L, Y141C stained for endogenous KDM2A (anti-KDM2A) and exogenous KDM2A (anti-HA). Tubulin was used as loading control. (h-i) WB quantification of endogenous KDM2A (h) and exogenous KDM2A (i) in HEK cells (*p<0.05, N = 3). One-way ANOVA were preformed to determine significance in panel b, c, e, f, h, and i. All quantifications represent mean ± s.e.m.

Given the altered distribution of both exogenously expressed and endogenous KDM2A protein caused by P235L, we next sought to assess the impact of these variants on KDM2A protein levels. Specifically, we introduced WT, P235L, and Y141C plasmids into HEK cells and examined both exogenous and endogenous KDM2A through Western blotting (Figure 2G). In the WT and Y141C conditions, there was no notable change in the expression of endogenous KDM2A. However, the P235L variant significantly decreased the levels of both exogenously expressed and endogenous KDM2A proteins (Figure 2G-I). This suggests that the P235L variants likely disrupt the stability of KDM2A proteins and exert a dominant-negative effect. To investigate the mechanisms underlying the reduced KDM2A levels, we compared the stability of KDM2A between the WT and P235L mutant protein. We performed Western blot (WB) analysis on HEK cells transfected with either the WT or P235L plasmid. Protein lysates were harvested at 0, 6, 12, 24, and 48 hours after cycloheximide (CHX) treatment (Figure 3A, 3B). The WB analysis revealed that KDM2A protein is less stable in the P235L variant-expressing cells, with a half-life (t_1/2_) of 4.35 hours compared to a t_1/2_ of 7.04 hours in WT-expressing cells. This suggests that the differential reduction of KDM2A in P235L variants is due to differences in the stability of the KDM2A protein.

**Figure 3.**
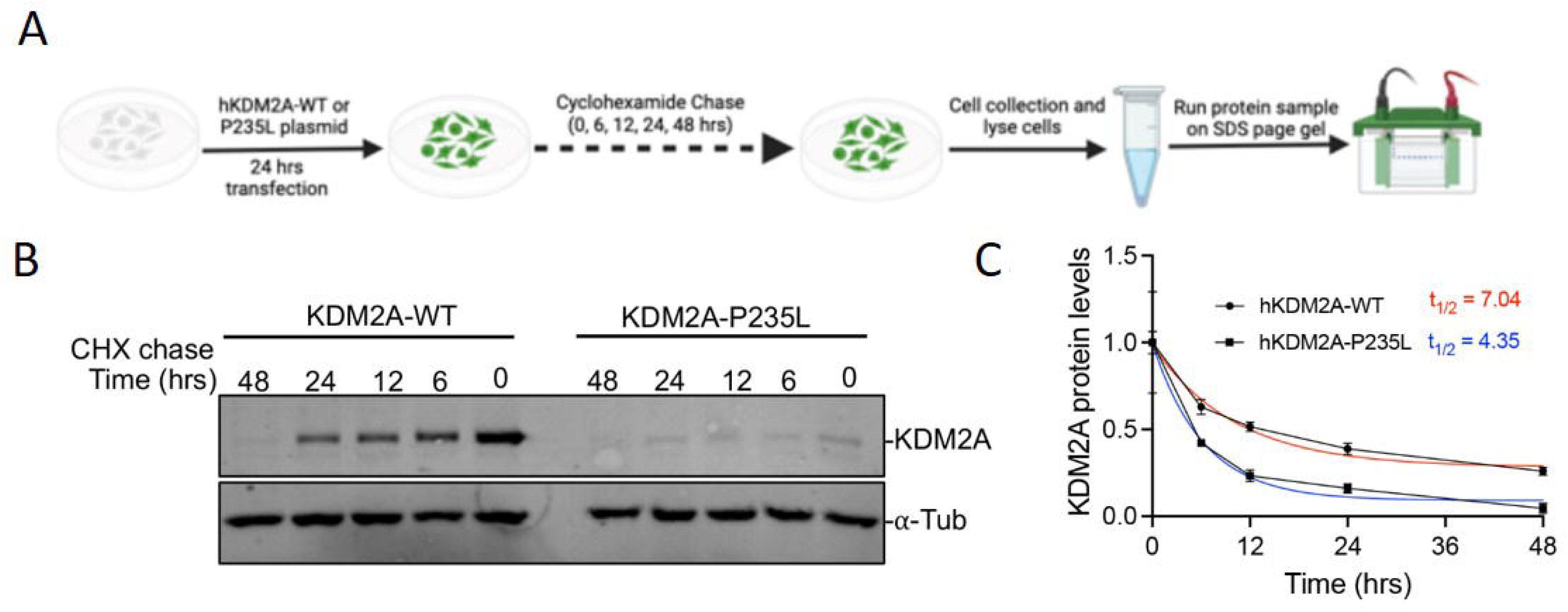
The P235L mutation alters the stability of the KDM2A protein. (a) Schematic of the cycloheximide (CHX) experiment conducted in HEK293T cells. (b) Representative Western blot showing KDM2A protein levels (anti-HA) in HEK293T cells transfected with either WT or P235L KDM2A plasmid at 0, 6, 12, 24, and 48 hours following CHX addition. Tubulin was used as a normalization control. (c) Quantification of the degradation rate and half-life (t_1/2_) of KDM2A after CHX treatment revealed an accelerated depletion of KDM2A in P235L-expressing cells compared to exogenous WT expression (nonlinear regression—one-phase decay, n=3).

### KDM2A variants cause eye degeneration, motor defects, and reduce lifespan in a Drosophila model

We next investigated the *in vivo* effects of missense variants in the Drosophila model system. To achieve this, we generated transgenic Drosophila expressing human KDM2A WT and missense variants (P235L, Y141C, and H811N) through PhiC31 integrase-mediated site-specific insertion of a single copy of the human *KDM2A* gene. The expression of these transgenes in flies using the eye tissues-targeting GMR-gl4 driver revealed a mutation-dependent rough eye phenotype, with the P235L variant exhibiting more pronounced effects (Figure 4A, 4B). The expression of WT showed a very mild phenotype. Likewise, in line with with observations from HEK 293T cells, the expression of the P235L variant in fly eyes resulted in a notable decrease in KDM2A protein levels (Figure 4C, 4D).

**Figure 4.**
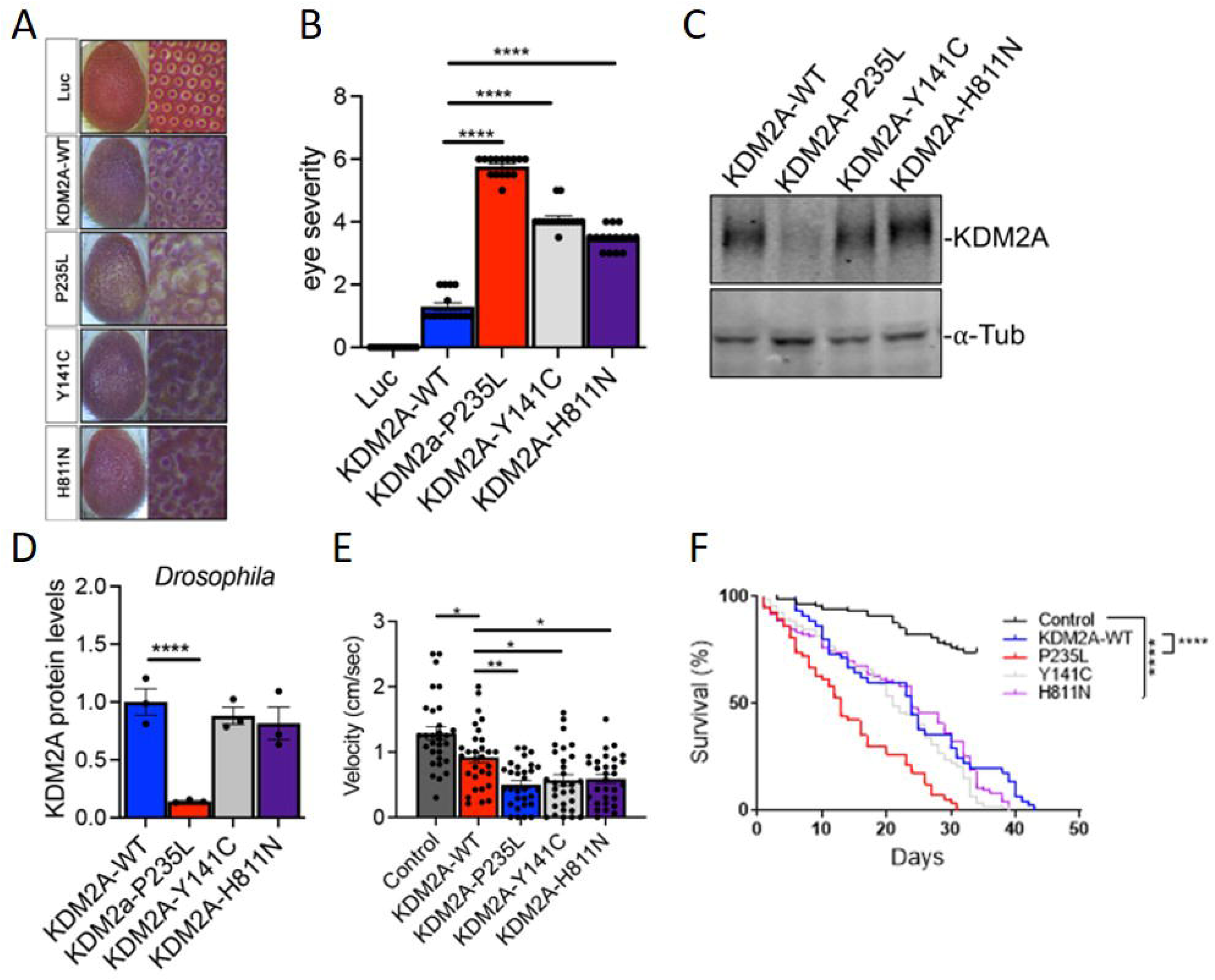
A Drosophila model expressing KDM2A variants display differential toxicity *in vivo*. (a) Representative panel of adult Drosophila eyes expressing KDM2A-WT, P235L, Y141C, H811N or luciferase (luc) control. (b) Quantification of eye degeneration severity demonstrated that KDM2A variants significantly enhance toxicity as compared to wild type KDM2A (****p<0.0001, N=15-20). (c) WB of Drosophila expressing KDM2A (WT, P235L, Y141C, and H811N) in the eye (GMR-gal4) stained with anti-KDM2A and anti-tubulin. (d) WB quantification of KDM2A in Drosophila showed that P235L but not the Y141C or H811N variants had a significantly reduced protein level compared to KDM2A-WT (****p<0.0001, N = 3). (e) Quantification of climbing velocity (cm/s) in flies expressing KDM2A-WT, P235L, Y141C, and H811N pan-neuronally (Elav-gal4) compared to Luciferase control or wild type KDM2A (n = 3 trials, 10 animals per trials, **p < 0.01, *p < 0.05). (f) Kaplan-Meier survival curve of flies expressing KDM2A-WT, P235L, Y141C, and H811N in neurons compared with Luciferase control (n = 50–80, ****p < 0.0001). One-way ANOVA was performed in b, d, and c, while Log-rank with Grehan-Breslow-Wilcoxon tests were performed to determine significance for panel f. All quantifications represent mean ± s.e.m.

To assess wether ectopic expression of KDM2A variants could cause motor deficits, we evaluated locomotion in 20-day-old flies. We expressed these variants pan-neuronally with the Elav-gal4 driver. While expression of KDM2A WT mildly but significantly reduced climbing velocity, a more profound decrease in climbing ability was observed in flies expressing the KDM2A mutants, particularly in the P235L variant (Figure 4E). To further determine the toxic impact of KDM2A, we assessed the effects of KDM2A variants on survival. Pan-neuronal expression of WT or mutant KDM2A significantly reduced survival compared to the luciferase control. However, the P235L variant led to a significantly shorter lifespan compared to both KDM2A WT or the other variants (Figure 4F), suggesting that the P235L variant exerts a more toxic effect *in vivo*.

Given our data showing that the P235L variant disrupts normal subcellular distribution and potentially impairs the function of endogenous KDM2A protein, we further investigated the toxicity of KDM2A variants in a Drosophila model where the endogenous Kdm2 gene (the single fly homolog of human KDM2A and KDM2B) is either knocked down (KD) or knocked out (KO). First, we knocked down Kdm2 in Drosophila eyes while expressing human wild-type (WT) and mutant (P235L, Y141C, and H811N) KDM2A. KD of endogenous Kdm2 in Drosophila eyes did not result in any overt degeneration, and expression of human KDM2A-WT in the Kdm2 KD background produced only a mild phenotype. In contrast, expression of the human KDM2A variants (P235L, Y141C, and H811N) in the Kdm2 KD background significantly exacerbated the degenerative eye phenotype compared to their respective controls (Figure 5A, 5B).

**Figure 5.**
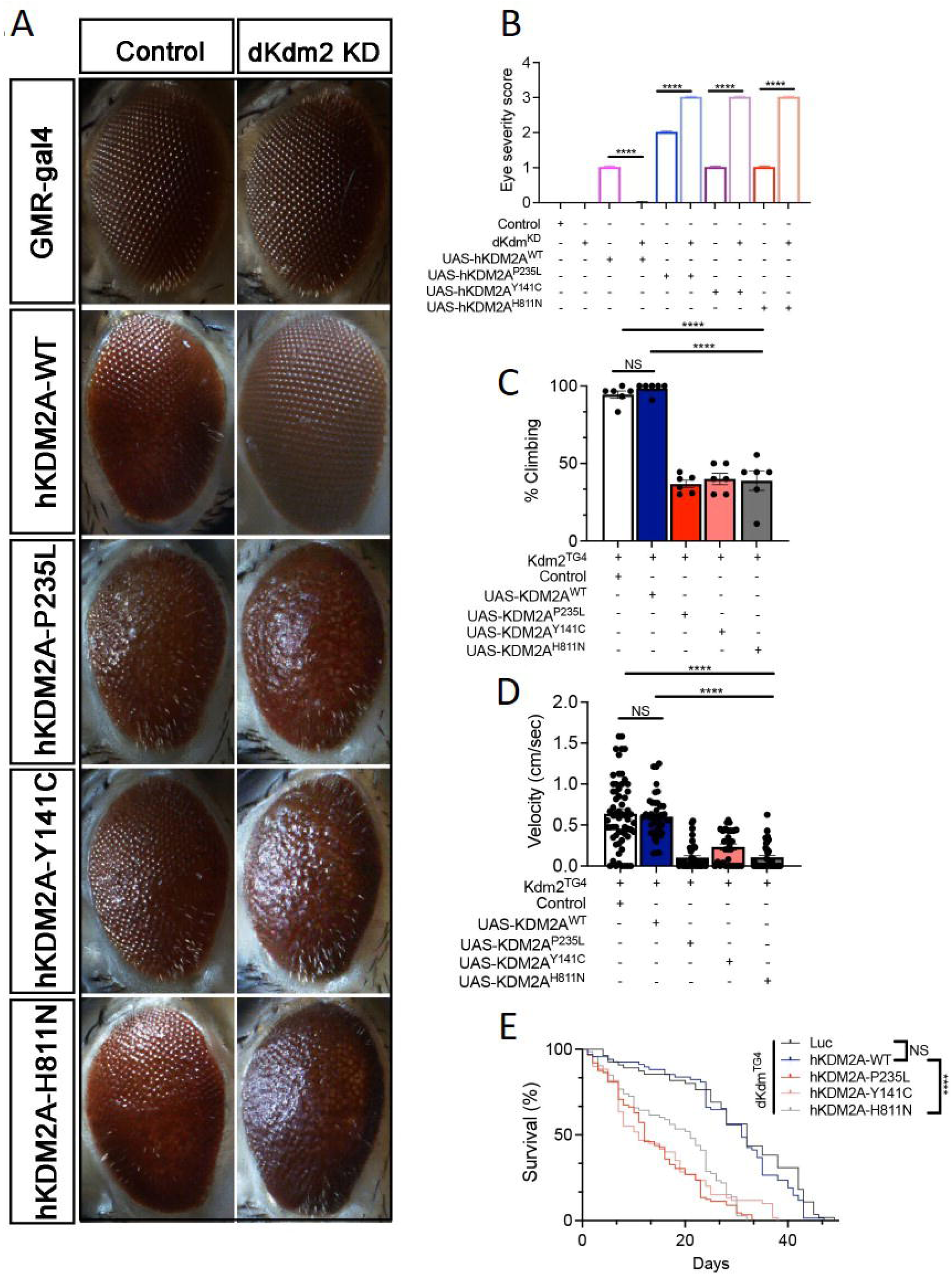
Expression of human KDM2A variants in a Drosophila Kdm2 knockdown or knockout model increases toxicity. (a) Images of Drosophila eyes expressing human WT or mutant KDM2A in the context of either endogenous Kdm2 or RNAi-mediated Kdm2 knockdown (KD). (b) Quantification shows that human KDM2A variants significantly increase eye degeneration severity in the Kdm2 KD background compared to controls with endogenous Kdm2 (****p<0.0001, n=20). Notably, neither hKDM2A-WT with endogenous Kdm2 KD nor Kdm2 KD alone caused overt eye degeneration. (c-e) Quantification analyses revealed a significant reduction in (c) the percentage of flies capable of climbing (****p<0.0001, n=6 trials/10 flies per trial), (d) climbing velocity (****p<0.0001, n=3 trials/15-20 flies per trial), and (e) lifespan (Kaplan-Meier survival curve, ****p<0.0001, n=80-90 flies) in Kdm2 KO Drosophila expressing human KDM2A variants (P235L, Y141C, and H811N) using the Trojan-Gal4 system, compared to hKDM2A-WT with endogenous Kdm2 KD or KO flies alone. Statistical significance was determined by one-way ANOVA for panels b, c, and d, and by Log-rank with Grehan-Breslow-Wilcoxon tests for panel e. All data are presented as mean ± s.e.m.

We further utilized the Trojan-MiMIC Gal4 driver system, which employs Recombination-Mediated Cassette Exchange (RMCE) of a Mi{MIC} insertion, resulting in the expression of GAL4 under the control of Kdm2 regulatory sequences.^32,33^ Consequently, Drosophila Kdm2 is knocked out in the Trojan-Gal4Kdm2 lines, allowing for a robust analysis of human KDM2A variants in a clean genetic background. We assessed motor function and lifespan in adult flies expressing human KDM2A WT and its variants in the specific cell types where endogenous Drosophila Kdm2 is normally expressed. The lack of phenotype in Kdm2 knockout animals suggests that the loss of Kdm2 in adults is not critical under the conditions tested (Figure 5C, 5D). Similarly, we found that expression of human KDM2A wild-type did not affect motor function or lifespan compared to Kdm2 knockout flies. However, the human KDM2A mutations (P235L, Y141C, and H811N) resulted in significantly worse motor function and survival compared to both the knockout and the human wild-type scenarios (Figure 5C, 5D). Altogether, these findings suggest that these variants in KDM2A are toxic, possibly due to a gain-of-function mechanism.

### Methylome analysis

As KDM2A is a component of the epigenetic machinery, it was hypthesized that disruption of KDM2A function by *de novo* variants would cause an abnormal methylation pattern in peripheral blood as a de facto functional read-out. Analysis of enzymatic-methylation sequencing data of twelve individuals with pLoF variants and missense variants compared to a control group, identified of 817 differentially methylated regions (DMRs). After applying stringent filtering criteria (p-adj<0.01 and minimum mean methylation difference > 10%), we defined a first episignature of the KDM2A cases. This signature includes 414 DMRs (406 hypermethylated and 8 hypomethylated regions) with a median methylation difference of 17.4%, and a median length of ∼200nts/16CpGs (Table S8). Hierarchical clustering of cases and controls revealed a distinct KDM2A cluster with individuals harboring frameshift variants (individuals 14, 15, 16 and 18) and missense variants (individuals 2, 4, 7 and 8) grouping together within this KDMA2 cluster (Figure 6). Data of four individuals (individuals 1, 5, 9 and 11) with missense variants was shown to group outside the bigger KDMA2 cluster, suggesting that these variants may not cause a severe disruption of KDM2A function (Figure S3).

**Figure 6.**
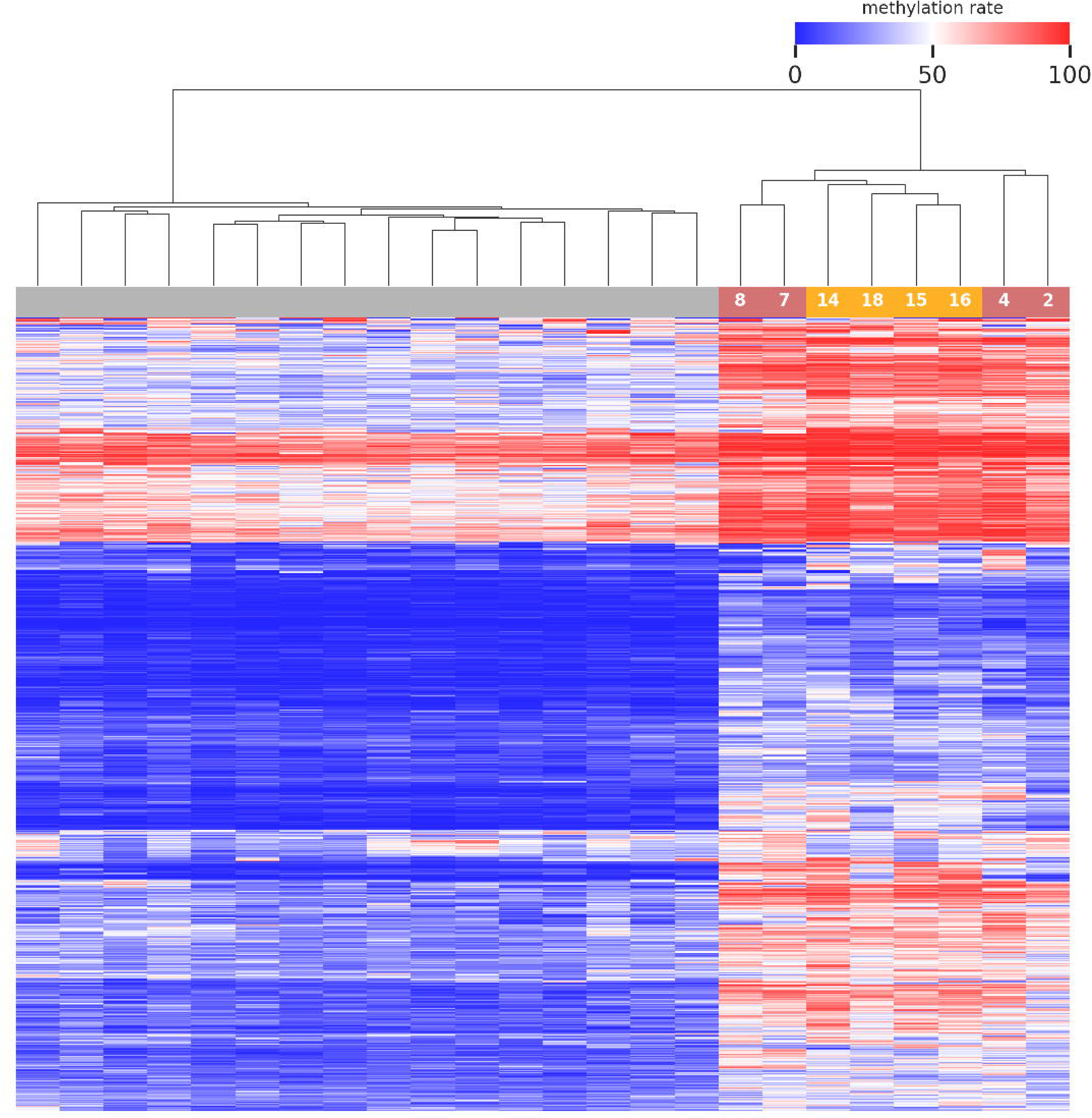
Episignature of pLoF and missense variants in *KDM2A.* Heatmap displays hierarchical clustering of selected CpG sites of the episignature. Columns represent probes (grey: control probes; yellow: *KDM2A* pLoF variants; light red: *KDM2A* missense variants; number represents individual in the cohort). Rows represent CpG sites. Color represents methylation ranging from dark blue (no methylation) to dark red (full methylation). A distinct separation between control samples and those from individuals with variants in *KMD2A* is observed.

## Discussion

In this study, we provide a detailed description of 18 individuals with heterozygous variants in *KDM2A*, including 17 with confirmed *de novo* status, all presenting with a syndromic neurodevelopmental disorder.

The affected individuals show an overlapping phenotype of DD/ID and several recurrent symptoms of note: growth abnormalities (including IUGR and short stature), microcephaly and feeding difficulties that are present in 35 – 72 % of individuals. DD/ID and growth abnormalities are a recurrent symptom among the Mendelian disorders of the epigenetic machinery,^3^ underscoring that the *KDM2A*-related rare disease delineated here shows a clinical overlap. Many of these disorders also present with a recognizable facial gestalt and include some of the classic dysmorphic syndromes such as Kabuki syndrome (e.g. *KMT2D*, MIM 147920) or Rubinstein-Taybi syndrome (e.g. *CREBBP*, MIM 180849). The affected individuals in the *KDM2A* cohort also showed facial dysmorphism, highlighted by recurrent observations of epicanthus, upslanted palpebral fissures, thin upper and/or lower lips and low-set ears as potentially defining features (Figure 1B). Deep phenotyping of facial images of affected individuals with the help of facial recognition tools could help to detect this potential typical facial gestalt of the *KDM2A*-related syndromic neurodevelopmental disorder in routine clinical genetic care.^34,35^ A malformation of cortical development (MCD), like the polymicrogyria observed in individual 4 is not a phenotype seen in other disorders of the epigenetic machinery. The trio exome analysis of this individual did not reveal any additional causative variant(s) for this phenotype in genes known to cause MCD nor additional variant(s) in other candidate genes.^36^ It remains therefore unclear, wether polymicrogyria is part of the phenotypic spectrum of the *KDM2A*-related disorder or if another, undetected genetic cause is involved. Of note, feeding difficulties were only present in individuals with *de novo* missense variants in *KDM2A*, and absent in individuals with pLoF variants suggesting a potential genotype-phenotype correlation. Similarly, microcephaly was observed exclusively in individuals with missense variants, with the exception of the prenatal case carrying a pLoF variant. Other recurrent phenotypes, however, did not show any potential genotype-phenotype correlations as they were observed in individuals that harbor missense variant and individuals with pLoF variants.

According to the data from gnomAD (v4 dataset),^14^ *KDM2A* is a gene with a significantly reduced number of pLoF and missense variants. This indicates a selective constraint on both types of variants in a control population that lacks severe, early-onset phenotypes such as DD, ID, microcephaly or short stature (missense o/e = 0.49, z-score 6.9; pLoF o/e = 0.02, LOEUF = 0.06). Among KDM genes (lysine demethylases), *KDM2A* is the most highly constrained gene with respect to both missense variants (Z-score in gnomAD) and pLOF variants (LOEUF, see constraint score landscape of KDM genes in Figure S1 as well as Table S6).

It is now also possible to evaluate prior assessments on what phenotypes could be caused by variants in candidate genes such as *KDM2A*. In a recent work by Dhindsa *et al.*, using a machine learning approach based on gene constraint, expression and many other gene-level annotations, *KDM2A* was predicted to cause an autosomal dominant phenotype of DD, developmental epileptic encephalopathy and autism on the 99.5 percentile or higher for each phenotype individually.^37^ This prior assessment underscores our phenotypic findings, including that both autism and epilepsy are important parts of the phenotypic spectrum of the *KDM2A*-related syndromic disorder, although only five affected individuals presented with seizures and four had a diagnosis of autism.

The identified *de novo* missense variants in *KDM2A* are located in or around the JmjC- and CxxC-domain, essential for the demethylation activity and DNA binding, respectively.^1,38^ In *KDM2B*, *de novo* missense variants are also preferably located in or around these two domains specifically (Figure S2).

In this study, we investigated the consequence of missense variants in KDM2A on the subcellular distribution of the KDM2A protein and toxicity. Previous studies demonstrated that KDM2A protein primarily localizes to the nucleus, where it binds to unmethylated CpG DNA via the ZF-CxxC domain.^30^ Consistent with these findings, KDM2A WT expressed in HEK293T cells exhibited predominantly nuclear localization. In contrast, our observations revealed distinct localization patterns for the Y141C and H811N variants, which displayed primarily nuclear with occasional cytoplasmic aggregation, while the P235L variant showed significant nuclear exclusion and cytoplasmic accumulation (Figure 2). Moreover, the endogenously expressed KDM2A protein undergoes cytoplasmic redistribution in the presence of exogenously expressed P235L variant. This observation was further supported by nuclear-cytoplasmic fractionation experiments, which consistently demonstrated predominant cytoplasmic localization of the P235L variant, suggesting a potential disruption in nuclear import. The loss of nuclear KDM2A protein may impair its ability to repress the transcription of centromeric satellite repeats and maintaining a heterochromatic state.^31^ Additionally, nuclear redistribution of KDM2A to the cytoplasm may also impair its ability to bind and regulate the stability of non-phosphorylated nuclear β-catenin, thereby potentially affecting the Wnt/β-catenin signaling pathway.^39^ Alternatively, cytoplasmic accumulation of KDM2A protein may confer a toxic gain-of-function. However, further investigations are required to elucidate the precise underlying mechanism. Moreover, our data revealed that the forced expression of the P235L variant is accompanied by a significant reduction in both endogenous and exogenously expressed KDM2A protein levels indicating compromised protein stability. This was confirmed through cycloheximide chase experiments which showed that the P235L variant reduces the half-life of KDM2A protein, suggesting that this variant negatively impacts protein stability.

The observed redistribution of KDM2A from the nucleus to the cytoplasm may exert toxic effects on cells. Similar mechanisms have been reported in amyotrophic lateral sclerosis and frontotemporal dementia (ALS/FTD) where pathogenic variants in proteins like FUS and TDP-43, lead to nuclear clearance contributing to cellular toxicity in various animal models including flies.^23,40^ To test whether KDM2A variants induce toxicity in an *in vivo* system, we generated Drosophila models expressing three missense variants (Y141C, P235L, and H811N). Ectopic expression of KDM2A in neuronal cells led to eye degeneration, motor deficits, and reduced survival (Figure 2), with the P235L variant exhibiting the highest toxicity, consistent with our *in vitro* findings. While KDM2A WT caused mild toxicity, this parallels observations from FUS expression studies in flies.^23^ Our data suggests both loss of nuclear function or and a potential gain-of-function effect in the cytoplasm contribute to the observed cellular toxicity, particularly evident in the P235L variant. Further research is needed to examine the exact mechanisms underlying KDM2A variant-mediated toxicity. Our *in vivo* fly data using a Kdm knockout (KO) or knockdown (KD) system suggested a possible dominant-negative effect of KDM2A variants. Our data showed that the expression of human wild-type KDM2A in the KD or KO background did not affect eye severity, motor function, or lifespan, which were similar to those observed in KD or KO Kdm flies. In contrast, expression of KDM2A variants resulted in significantly worse phenotypes compared to both KO flies and those expressing human KDM2A WT. These findings suggest that variants in KDM2A may confer a new or enhanced toxic effect, characteristic of gain-of-function mutations.

We analysed blood derived DNA and detected a *KDM2A*-related episignature, further underscoring a shared pathomechanism among affected individuals. In the future, with further refinement, this episignature could serve as a valuable tool for interpreting variants of unknown significance and aid in the diagnosis of affected individuals.

In summary, we use different lines of clinical, genetic, functional and epigenetic evidence to firmly establish *de novo* variants in *KDM2A* as the cause of a syndromic neurodevelopmental disorder.

## Data and code availability

All identified variants in *KDM2A* have been uploaded to ClinVar https://www.ncbi.nlm.nih.gov/clinvar/submitters/506086/.

FastQC is availabe at https://www.bioinformatics.babraham.ac.uk/projects/fastqc/

BWA-meth is available at https://github.com/brentp/bwa-meth.

Picard is available at https://broadinstitute.github.io/picard/

MethylDackel is available at https://github.com/dpryan79/MethylDackel

Pipeline for enzymatic-methylation sequencing data processing is available at: https://github.com/StephanHolgerD/methylmappdedupmethyldackelsnakemake

## Supplemental information

Supplemental data includes detailed case reports of the described individuals, two tables (Table S1, Table S2). Tables S3-S7 are provided separately as an Excel file.

## Consortia

The members of the Undiagnosed Diseases Network are Maria T. Acosta, David R. Adams, Ben Afzali, Ali Al-Beshri, Eric Allenspach, Aimee Allworth, Raquel L. Alvarez, Justin Alvey, Ashley Andrews, Euan A. Ashley, Carlos A. Bacino, Guney Bademci, Ashok Balasubramanyam, Dustin Baldridge, Jim Bale, Michael Bamshad, Deborah Barbouth, Pinar Bayrak-Toydemir, Anita Beck, Alan H. Beggs, Edward Behrens, Gill Bejerano, Hugo J. Bellen, Jimmy Bennett, Jonathan A. Bernstein, Gerard T. Berry, Anna Bican, Stephanie Bivona, Elizabeth Blue, John Bohnsack, Devon Bonner, Nicholas Borja, Lorenzo Botto, Lauren C. Briere, Elizabeth A. Burke, Lindsay C. Burrage, Manish J. Butte, Peter Byers, William E. Byrd, Kaitlin Callaway, John Carey, George Carvalho, Thomas Cassini, Sirisak Chanprasert, Hsiao-Tuan Chao, Ivan Chinn, Gary D. Clark, Terra R. Coakley, Laurel A. Cobban, Joy D. Cogan, Matthew Coggins, F. Sessions Cole, Brian Corner, Rosario I. Corona, William J. Craigen, Andrew B. Crouse, Vishnu Cuddapah, Precilla D’Souza, Hongzheng Dai, Kahlen Darr, Surendra Dasari, Joie Davis, Margaret Delgado, Esteban C. Dell’Angelica, Katrina Dipple, Daniel Doherty, Naghmeh Dorrani, Jessica Douglas, Emilie D. Douine, Dawn Earl, Lisa T. Emrick, Christine M. Eng, Cecilia Esteves, Kimberly Ezell, Elizabeth L. Fieg, Paul G. Fisher, Brent L. Fogel, Jiayu Fu, William A. Gahl, Rebecca Ganetzky, Emily Glanton, Ian Glass, Page C. Goddard, Joanna M. Gonzalez, Andrea Gropman, Meghan C. Halley, Rizwan Hamid, Neal Hanchard, Kelly Hassey, Nichole Hayes, Frances High, Anne Hing, Fuki M. Hisama, Ingrid A. Holm, Jason Hom, Martha Horike-Pyne, Alden Huang, Yan Huang, Anna Hurst, Wendy Introne, Gail P. Jarvik, Suman Jayadev, Orpa Jean-Marie, Vaidehi Jobanputra, Oguz Kanca, Yigit Karasozen, Shamika Ketkar, Dana Kiley, Gonench Kilich, Eric Klee, Shilpa N. Kobren, Isaac S. Kohane, Jennefer N. Kohler, Bruce Korf, Susan Korrick, Deborah Krakow, Elijah Kravets, Seema R. Lalani, Christina Lam, Brendan C. Lanpher, Ian R. Lanza, Kumarie Latchman, Kimberly LeBlanc, Brendan H. Lee, Kathleen A. Leppig, Richard A. Lewis, Pengfei Liu, Nicola Longo, Joseph Loscalzo, Richard L. Maas, Ellen F. Macnamara, Calum A. MacRae, Valerie V. Maduro, AudreyStephannie Maghiro, Rachel Mahoney, May Christine V. Malicdan, Rong Mao, Ronit Marom, Gabor Marth, Beth A. Martin, Martin G. Martin, Julian A. Martínez-Agosto, Shruti Marwaha, Allyn McConkie-Rosell, Ashley McMinn, Matthew Might, Mohamad Mikati, Danny Miller, Ghayda Mirzaa, Breanna Mitchell, Paolo Moretti, Marie Morimoto, John J. Mulvihill, Lindsay Mulvihill, Mariko Nakano-Okuno, Stanley F. Nelson, Serena Neumann, Dargie Nitsuh, Donna Novacic, Devin Oglesbee, James P. Orengo, Laura Pace, Stephen Pak, J. Carl Pallais, Neil H. Parker, LéShon Peart, Leoyklang Petcharet, John A. Phillips III, Filippo Pinto e Vairo, Jennifer E. Posey, Lorraine Potocki, Barbara N. Pusey Swerdzewski, Aaron Quinlan, Daniel J. Rader, Ramakrishnan Rajagopalan, Deepak A. Rao, Anna Raper, Wendy Raskind, Adriana Rebelo, Chloe M. Reuter, Lynette Rives, Lance H. Rodan, Martin Rodriguez, Jill A. Rosenfeld, Elizabeth Rosenthal, Francis Rossignol, Maura Ruzhnikov, Marla Sabaii, Jacinda B. Sampson, Timothy Schedl, Lisa Schimmenti, Kelly Schoch, Daryl A. Scott, Elaine Seto, Vandana Shashi, Emily Shelkowitz, Sam Sheppeard, Jimann Shin, Edwin K. Silverman, Giorgio Sirugo, Kathy Sisco, Tammi Skelton, Cara Skraban, Carson A. Smith, Kevin S. Smith, Lilianna Solnica-Krezel, Ben Solomon, Rebecca C. Spillmann, Andrew Stergachis, Joan M. Stoler, Kathleen Sullivan, Shamil R. Sunyaev, Shirley Sutton, David A. Sweetser, Virginia Sybert, Holly K. Tabor, Queenie Tan, Arjun Tarakad, Herman Taylor, Mustafa Tekin, Willa Thorson, Cynthia J. Tifft, Camilo Toro, Alyssa A. Tran, Rachel A. Ungar, Adeline Vanderver, Matt Velinder, Dave Viskochil, Tiphanie P. Vogel, Colleen E. Wahl, Melissa Walker, Nicole M. Walley, Jennifer Wambach, Michael F. Wangler, Patricia A. Ward, Daniel Wegner, Monika Weisz Hubshman, Mark Wener, Tara Wenger, Monte Westerfield, Matthew T. Wheeler, Jordan Whitlock, Lynne A. Wolfe, Heidi Wood, Kim Worley, Shinya Yamamoto, Zhe Zhang and Stephan Zuchner.

## Data Availability

All data produced in the present study are available upon reasonable request to the authors.

## Acknowledgements

We thank all families who participated in this study and generously contributed their time and data. SS is funded through Albert Rowe II endowed chair in Genetics. Research reported in this manuscript was in part supported by the NIH Common Fund, through the Office of Strategic Coordination/Office of the NIH Director and the National Institute of Neurological Disorders and Stroke of the NIH under Award Numbers U01HG010218 and U01HG007708. The content is solely the responsibility of the authors and does not necessarily represent the official views of the National Institutes of Health.

## Web resources

GenBank, https://www.ncbi.nlm.nih.gov/genbank/

OMIM, https://www.omim.org/

GeneMatcher, https://genematcher.org/

gnomAD, https://gnomad.broadinstitute.org/

## References

1. Kawakami, E., Tokunaga, A., Ozawa, M., Sakamoto, R., and Yoshida, N. (2015). The histone demethylase Fbxl11/Kdm2a plays an essential role in embryonic development by repressing cell-cycle regulators. Mechanisms of Development 135, 31–42. 10.1016/j.mod.2014.10.001.

2. Levy, M.A., McConkey, H., Kerkhof, J., Barat-Houari, M., Bargiacchi, S., Biamino, E., Bralo, M.P., Cappuccio, G., Ciolfi, A., Clarke, A., et al. (2022). Novel diagnostic DNA methylation episignatures expand and refine the epigenetic landscapes of Mendelian disorders. Human Genetics and Genomics Advances 3, 100075. 10.1016/j.xhgg.2021.100075.

3. Harris, J.R., Gao, C.W., Britton, J.F., Applegate, C.D., Bjornsson, H.T., and Fahrner, J.A. (2023). Five years of experience in the Epigenetics and Chromatin Clinic: what have we learned and where do we go from here? Hum Genet. 10.1007/s00439-023-02537-1.

4. Faundes, V., Newman, W.G., Bernardini, L., Canham, N., Clayton-Smith, J., Dallapiccola, B., Davies, S.J., Demos, M.K., Goldman, A., Gill, H., et al. (2018). Histone Lysine Methylases and Demethylases in the Landscape of Human Developmental Disorders. The American Journal of Human Genetics 102, 175–187. 10.1016/j.ajhg.2017.11.013.

5. van Jaarsveld, R.H., Reilly, J., Cornips, M.-C., Hadders, M.A., Agolini, E., Ahimaz, P., Anyane-Yeboa, K., Bellanger, S.A., van Binsbergen, E., van den Boogaard, M.-J., et al. (2022). Delineation of a KDM2B-related neurodevelopmental disorder and its associated DNA methylation signature. Genetics in Medicine, S109836002200942X. 10.1016/j.gim.2022.09.006.

6. Rots, D., Jakub, T.E., Keung, C., Jackson, A., Banka, S., Pfundt, R., De Vries, B.B.A., Van Jaarsveld, R.H., Hopman, S.M.J., Van Binsbergen, E., et al. (2023). The clinical and molecular spectrum of the KDM6B-related neurodevelopmental disorder. The American Journal of Human Genetics 110, 963–978. 10.1016/j.ajhg.2023.04.008.

7. Kosmicki, J.A., Samocha, K.E., Howrigan, D.P., Sanders, S.J., Slowikowski, K., Lek, M., Karczewski, K.J., Cutler, D.J., Devlin, B., Roeder, K., et al. (2017). Refining the role of de novo protein-truncating variants in neurodevelopmental disorders by using population reference samples. Nat Genet 49, 504–510. 10.1038/ng.3789.

8. Zhou, X., Feliciano, P., Shu, C., Wang, T., Astrovskaya, I., Hall, J.B., Obiajulu, J.U., Wright, J.R., Murali, S.C., Xu, S.X., et al. (2022). Integrating de novo and inherited variants in 42,607 autism cases identifies mutations in new moderate-risk genes. Nat Genet 54, 1305–1319. 10.1038/s41588-022-01148-2.

9. Fu, J.M., Satterstrom, F.K., Peng, M., Brand, H., Collins, R.L., Dong, S., Wamsley, B., Klei, L., Wang, L., Hao, S.P., et al. (2022). Rare coding variation provides insight into the genetic architecture and phenotypic context of autism. Nat Genet 54, 1320–1331. 10.1038/s41588-022-01104-0.

10. Damianidou, E., Mouratidou, L., and Kyrousi, C. (2022). Research models of neurodevelopmental disorders: The right model in the right place. Front Neurosci 16, 1031075. 10.3389/fnins.2022.1031075.

11. Kour, S., Rajan, D.S., Fortuna, T.R., Anderson, E.N., Ward, C., Lee, Y., Lee, S., Shin, Y.B., Chae, J.-H., Choi, M., et al. (2021). Loss of function mutations in GEMIN5 cause a neurodevelopmental disorder. Nat Commun 12, 2558. 10.1038/s41467-021-22627-w.

12. García-Cazorla, À., Verdura, E., Juliá-Palacios, N., Anderson, E.N., Goicoechea, L., Planas-Serra, L., Tsogtbaatar, E., Dsouza, N.R., Schlüter, A., Urreizti, R., et al. (2020). Impairment of the mitochondrial one-carbon metabolism enzyme SHMT2 causes a novel brain and heart developmental syndrome. Acta Neuropathol 140, 971–975. 10.1007/s00401-020-02223-w.

13. Sobreira, N., Schiettecatte, F., Valle, D., and Hamosh, A. (2015). GeneMatcher: a matching tool for connecting investigators with an interest in the same gene. Hum Mutat 36, 928–930. 10.1002/humu.22844.

14. Chen, S., Francioli, L.C., Goodrich, J.K., Collins, R.L., Kanai, M., Wang, Q., Alföldi, J., Watts, N.A., Vittal, C., Gauthier, L.D., et al. (2024). A genomic mutational constraint map using variation in 76,156 human genomes. Nature 625, 92–100. 10.1038/s41586-023-06045-0.

15. Morales, J., Pujar, S., Loveland, J.E., Astashyn, A., Bennett, R., Berry, A., Cox, E., Davidson, C., Ermolaeva, O., Farrell, C.M., et al. (2022). A joint NCBI and EMBL-EBI transcript set for clinical genomics and research. Nature 604, 310–315. 10.1038/s41586-022-04558-8.

16. Richards, S., Aziz, N., Bale, S., Bick, D., Das, S., Gastier-Foster, J., Grody, W.W., Hegde, M., Lyon, E., Spector, E., et al. (2015). Standards and guidelines for the interpretation of sequence variants: a joint consensus recommendation of the American College of Medical Genetics and Genomics and the Association for Molecular Pathology. Genetics in Medicine 17, 405–424. 10.1038/gim.2015.30.

17. Rentzsch, P., Schubach, M., Shendure, J., and Kircher, M. (2021). CADD-Splice—improving genome-wide variant effect prediction using deep learning-derived splice scores. Genome Med 13, 31. 10.1186/s13073-021-00835-9.

18. Ioannidis, N.M., Rothstein, J.H., Pejaver, V., Middha, S., McDonnell, S.K., Baheti, S., Musolf, A., Li, Q., Holzinger, E., Karyadi, D., et al. (2016). REVEL: An Ensemble Method for Predicting the Pathogenicity of Rare Missense Variants. The American Journal of Human Genetics 99, 877–885. 10.1016/j.ajhg.2016.08.016.

19. Pejaver, V., Urresti, J., Lugo-Martinez, J., Pagel, K.A., Lin, G.N., Nam, H.-J., Mort, M., Cooper, D.N., Sebat, J., Iakoucheva, L.M., et al. (2020). Inferring the molecular and phenotypic impact of amino acid variants with MutPred2. Nat Commun 11, 5918. 10.1038/s41467-020-19669-x.

20. Carter, H., Douville, C., Stenson, P.D., Cooper, D.N., and Karchin, R. (2013). Identifying Mendelian disease genes with the Variant Effect Scoring Tool. BMC Genomics 14, S3. 10.1186/1471-2164-14-S3-S3.

21. Feng, B.-J. (2017). PERCH: A Unified Framework for Disease Gene Prioritization. Human Mutation 38, 243–251. 10.1002/humu.23158.

22. Pejaver, V., Byrne, A.B., Feng, B.-J., Pagel, K.A., Mooney, S.D., Karchin, R., O’Donnell-Luria, A., Harrison, S.M., Tavtigian, S.V., Greenblatt, M.S., et al. (2022). Calibration of computational tools for missense variant pathogenicity classification and ClinGen recommendations for PP3/BP4 criteria. The American Journal of Human Genetics 109, 2163–2177. 10.1016/j.ajhg.2022.10.013.

23. Casci, I., Krishnamurthy, K., Kour, S., Tripathy, V., Ramesh, N., Anderson, E.N., Marrone, L., Grant, R.A., Oliver, S., Gochenaur, L., et al. (2019). Muscleblind acts as a modifier of FUS toxicity by modulating stress granule dynamics and SMN localization. Nat Commun 10, 5583. 10.1038/s41467-019-13383-z.

24. Anderson, E.N., Morera, A.A., Kour, S., Cherry, J.D., Ramesh, N., Gleixner, A., Schwartz, J.C., Ebmeier, C., Old, W., Donnelly, C.J., et al. (2021). Traumatic injury compromises nucleocytoplasmic transport and leads to TDP-43 pathology. eLife 10, e67587. 10.7554/eLife.67587.

25. Anderson, E.N., Gochenaur, L., Singh, A., Grant, R., Patel, K., Watkins, S., Wu, J.Y., and Pandey, U.B. (2018). Traumatic injury induces stress granule formation and enhances motor dysfunctions in ALS/FTD models. Human Molecular Genetics 27, 1366–1381. 10.1093/hmg/ddy047.

26. Pandey, U.B., Nie, Z., Batlevi, Y., McCray, B.A., Ritson, G.P., Nedelsky, N.B., Schwartz, S.L., DiProspero, N.A., Knight, M.A., Schuldiner, O., et al. (2007). HDAC6 rescues neurodegeneration and provides an essential link between autophagy and the UPS. Nature 447, 860–864. 10.1038/nature05853.

27. Martin, M. (2011). Cutadapt removes adapter sequences from high-throughput sequencing reads. EMBnet.journal 17, 10–12. 10.14806/ej.17.1.200.

28. Jühling, F., Kretzmer, H., Bernhart, S.H., Otto, C., Stadler, P.F., and Hoffmann, S. (2016). metilene: fast and sensitive calling of differentially methylated regions from bisulfite sequencing data. Genome Res 26, 256–262. 10.1101/gr.196394.115.

29. Waskom, M.L. (2021). seaborn: statistical data visualization. Journal of Open Source Software 6, 3021. 10.21105/joss.03021.

30. Blackledge, N.P., Zhou, J.C., Tolstorukov, M.Y., Farcas, A.M., Park, P.J., and Klose, R.J. (2010). CpG Islands Recruit a Histone H3 Lysine 36 Demethylase. Molecular Cell 38, 179–190. 10.1016/j.molcel.2010.04.009.

31. Frescas, D., Guardavaccaro, D., Kuchay, S.M., Kato, H., Poleshko, A., Basrur, V., Elenitoba-Johnson, K.S., Katz, R.A., and Pagano, M. (2008). KDM2A represses transcription of centromeric satellite repeats and maintains the heterochromatic state. Cell Cycle 7, 3539–3547. 10.4161/cc.7.22.7062.

32. Lee, P.-T., Zirin, J., Kanca, O., Lin, W.-W., Schulze, K.L., Li-Kroeger, D., Tao, R., Devereaux, C., Hu, Y., Chung, V., et al. (2018). A gene-specific T2A-GAL4 library for Drosophila. eLife 7, e35574. 10.7554/eLife.35574.

33. Marcogliese, P.C., Deal, S.L., Andrews, J., Harnish, J.M., Bhavana, V.H., Graves, H.K., Jangam, S., Luo, X., Liu, N., Bei, D., et al. (2022). Drosophila functional screening of de novo variants in autism uncovers damaging variants and facilitates discovery of rare neurodevelopmental diseases. Cell Reports 38, 110517. 10.1016/j.celrep.2022.110517.

34. Hsieh, T.-C., Bar-Haim, A., Moosa, S., Ehmke, N., Gripp, K.W., Pantel, J.T., Danyel, M., Mensah, M.A., Horn, D., Rosnev, S., et al. (2022). GestaltMatcher facilitates rare disease matching using facial phenotype descriptors. Nat Genet 54, 349–357. 10.1038/s41588-021-01010-x.

35. Dingemans, A.J.M., Hinne, M., Truijen, K.M.G., Goltstein, L., Van Reeuwijk, J., De Leeuw, N., Schuurs-Hoeijmakers, J., Pfundt, R., Diets, I.J., Den Hoed, J., et al. (2023). PhenoScore quantifies phenotypic variation for rare genetic diseases by combining facial analysis with other clinical features using a machine-learning framework. Nat Genet. 10.1038/s41588-023-01469-w.

36. Oegema, R., Barakat, T.S., Wilke, M., Stouffs, K., Amrom, D., Aronica, E., Bahi-Buisson, N., Conti, V., Fry, A.E., Geis, T., et al. (2020). International consensus recommendations on the diagnostic work-up for malformations of cortical development. Nat Rev Neurol 16, 618–635. 10.1038/s41582-020-0395-6.

37. Dhindsa, R.S., Weido, B., Dhindsa, J.S., Shetty, A.J., Sands, C., Petrovski, S., Vitsios, D., and Zoghbi, A.W. (2022). Genome-wide prediction of dominant and recessive neurodevelopmental disorder risk genes. bioRxiv. 10.1101/2022.11.21.517436.

38. Tsukada, Y., Fang, J., Erdjument-Bromage, H., Warren, M.E., Borchers, C.H., Tempst, P., and Zhang, Y. (2006). Histone demethylation by a family of JmjC domain-containing proteins. Nature 439, 811–816. 10.1038/nature04433.

39. Lu, L., Gao, Y., Zhang, Z., Cao, Q., Zhang, X., Zou, J., and Cao, Y. (2015). Kdm2a/b Lysine Demethylases Regulate Canonical Wnt Signaling by Modulating the Stability of Nuclear β-Catenin. Developmental Cell 33, 660–674. 10.1016/j.devcel.2015.04.006.

40. Yang, C., Qiao, T., Yu, J., Wang, H., Guo, Y., Salameh, J., Metterville, J., Parsi, S., Yusuf, I., Brown, R.H., et al. (2022). Low-level overexpression of wild type TDP-43 causes late-onset, progressive neurodegeneration and paralysis in mice. PLoS One 17, e0255710. 10.1371/journal.pone.0255710.

