## Supplemental data for "De novo variants in *KDM2A* cause a syndromic neurodevelopmental disorder"

### Content

Case reports of individuals with causative variants in *KDM2A* ... 3
Individual 1, c.422A>G, p.(Tyr141Cys), de novo ... 3
Individual 2, c.704C>T, p.(Pro235Leu), de novo ... 4
Individual 3, c.850C>T, p.(His284Tyr), de novo ... 5
Individual 4, c.956G>A, p.(Arg319Gln), de novo ... 6
Individual 5, c.1571T>G, p.(Phe524Cys), de novo ... 7
Individual 6, c.1703G>A, (p.Arg568Gln), de novo ... 8
Individual 7, c.1772T>C, p.(Met591Thr), de novo ... 9
Individual 8, c.1796G>C, p.(Arg599Pro), de novo ... 10
Individual 9, c.2327A>G, p.(Lys776Arg), de novo ... 11
Individual 10, c.2328G>T, p.(Lys776Asn), de novo ... 12
Individual 11, c.2431C>A, p.(His811Asn), de novo ... 13
Individual 12, c.58C>T, p.(Arg20*), de novo ... 14
Individual 13, c.579C>G, p.(Tyr193*), de novo ... 15
Individual 14, c.1676dup, p.(Ile560Aspfs*71), de novo ... 16
Individual 15, c.1677delG, p.(Ile560Leufs*32), de novo ... 17
Individual 16, c.2404dup, p.(Thr802Asnfs*49), heterozygous ... 18
Individual 17, c.2667delC, p.(Asp889Glufs*47), de novo ... 19
Individual 18, c.2809_2812dup, p.(Cys938*), de novo ... 20
Case report of an individual with a variant in *KDM2A* but insufficient evidence for causality ... 21
Individual S19, c.2323G>A, p.(Glu775Lys), heterozygous ... 21
Table S1. Variant information and classification according to ACMG criteria. ... 22
Table S2. *In silico* predition of missense variants in *KDM2A*. ... 23
Table S3. Detailed clinical data of individuals with causative variants in *KDM2A* ... 24
Table S4. Detailed clinical data of an individual with variant in *KDM2A* but insufficient evidence for causality ... 24
Table S5. Annotation and *in silico* scores of all missense variants in *KDM2A* ... 24
Table S6. List of KDM genes ... 24
Table S7. Reported variants in the literature in KDM genes ... 24
Table S8. Methylation data of the *KDM2A*-related Episignature ... 24
Figure S1. Constraint score landscape of KDM genes. ... 25
Figure S2. Overview of domain structure and reported variants in the literature in KDM genes. ... 26
Figure S3. Methylation data of all variants. ... 27
References ... 28

Case reports of individuals with causative variants in *KDM2A*

#### Individual 1, c.422A>G, p.(Tyr141Cys), de novo

Removed due to medRxiv policy

#### Individual 2, c.704C>T, p.(Pro235Leu), de novo

Removed due to medRxiv policy

#### Individual 3, c.850C>T, p.(His284Tyr), de novo

Removed due to medRxiv policy

#### Individual 4, c.956G>A, p.(Arg319Gln), de novo

Removed due to medRxiv policy

#### Individual 5, c.1571T>G, p.(Phe524Cys), de novo

Removed due to medRxiv policy

#### Individual 6, c.1703G>A, (p.Arg568Gln), de novo

Removed due to medRxiv policy

#### Individual 7, c.1772T>C, p.(Met591Thr), de novo

Removed due to medRxiv policy

#### Individual 8, c.1796G>C, p.(Arg599Pro), de novo

Removed due to medRxiv policy

#### Individual 9, c.2327A>G, p.(Lys776Arg), de novo

Removed due to medRxiv policy

#### Individual 10, c.2328G>T, p.(Lys776Asn), de novo

Removed due to medRxiv policy

#### Individual 11, c.2431C>A, p.(His811Asn), de novo

Removed due to medRxiv policy

#### Individual 12, c.58C>T, p.(Arg20*), de novo

Removed due to medRxiv policy

#### Individual 13, c.579C>G, p.(Tyr193*), de novo

Removed due to medRxiv policy

#### Individual 14, c.1676dup, p.(Ile560Aspfs*71), de novo

Removed due to medRxiv policy

#### Individual 15, c.1677delG, p.(Ile560Leufs*32), de novo

Removed due to medRxiv policy

#### Individual 16, c.2404dup, p.(Thr802Asnfs*49), heterozygous

Removed due to medRxiv policy

#### Individual 17, c.2667delC, p.(Asp889Glufs*47), de novo

Removed due to medRxiv policy

#### Individual 18, c.2809_2812dup, p.(Cys938*), de novo

Removed due to medRxiv policy

Case report of an individual with a variant in *KDM2A* but insufficient evidence for causality

#### Individual S19, c.2323G>A, p.(Glu775Lys), heterozygous

Removed due to medRxiv policy

| individual | Chr11: g.(hg38) | c. | p. | allelic state | origin | predicted effect | ACMG criteria[1] | classification |
|---|---|---|---|---|---|---|---|---|
| 1 | 67207624 | c.422A>G | p.(Tyr141Cys) | heterozygous | de novo | missense | PS2_MOD, PS3_MOD, PM2, PP2, PP3 | likely pathogenic |
| 2 | 67217747 | c.704C>T | p.(Pro235Leu) | heterozygous | de novo | missense | PS2_MOD, PS3_MOD , PM2, PP2, PP3 | likely pathogenic |
| 3 | 67219296 | c.850C>T | p.(His284Tyr) | heterozygous | de novo | missense | PS2_MOD, PM2, PP2, PP3 | likely pathogenic |
| 4 | 67219402 | c.956G>A | p.(Arg319Gln) | heterozygous | de novo | missense | PS2_MOD, PM, PP2, PP3 | likely pathogenic |
| 5 | 67245196 | c.1571T>G | p.(Phe524Cys) | heterozygous | de novo | missense | PS2_MOD, PM2, PP2 | uncertain significance* |
| 6 | 67245328 | c.1703G>A | p.(Arg568Gln) | heterozygous | de novo | missense | PS2_MOD, PM2, PP2, PP3 | likely pathogenic |
| 7 | 67245397 | c.1772T>C | p.(Met591Thr) | heterozygous | de novo | missense | PS2_MOD, PM2, PP2, PP3 | likely pathogenic |
| 8 | 67245421 | c.1796G>C | p.(Arg599Pro) | heterozygous | de novo | missense | PS2_MOD, PM2, PP2, PP3 | likely pathogenic |
| 9 | 67250357 | c.2327A>G | p.(Lys776Arg) | heterozygous | de novo | missense | PS2_MOD, PM2, PP2 | uncertain significance* |
| 10 | 67250358 | c.2328G>T | p.(Lys776Asn) | heterozygous | de novo | missense | PS2_MOD, PM2, PP2 | uncertain significance* |
| 11 | 67250461 | c.2431C>A | p.(His811Asn) | heterozygous | de novo | missense | PS2_MOD, PS3_MOD, PM2, PP2 | likely pathogenic |
| 12 | 67180094 | c.58C>T | p.(Arg20*) | heterozygous | de novo | nonsense mediated mRNA decay | PVS1, PS2_MOD, PM2P | pathogenic |
| 13 | 66982903 | c.579C>G | p.(Tyr193*) | heterozygous | de novo | nonsense mediated mRNA decay | PVS1, PS2_MOD, PM2 | pathogenic |
| 14 | 67245301dup | c.1676dup | p.(Ile560Aspfs*71) | heterozygous | de novo | nonsense mediated mRNA decay | PVS1, PS2_MOD, PM2 | pathogenic |
| 15 | 67245302del | c.1677del | p.(Ile560Leufs*32) | heterozygous | de novo | nonsense mediated mRNA decay | PVS1, PS2_MOD, PM2 | pathogenic |
| 16 | 67250434dup | c.2404dup | p.(Thr802Asnfs*49) | heterozygous | unknown | nonsense mediated mRNA decay | PVS1, PM2 | likely pathogenic |
| 17 | 67250697del | c.2667del | p.(Asp889Glufs*47) | heterozygous | de novo | nonsense mediated mRNA decay | PVS1, PS2_MOD, PM2 | pathogenic |
| 18 | 67252734_ 67252737dup | c.2809_ 2812dup | p.(Cys938*) | heterozygous | de novo | nonsense mediated mRNA decay | PVS1, PS2_MOD, PM2 | pathogenic |

Table S1. Variant information and classification according to ACMG criteria.[1] MANE Select transcript NM_012308.3 was used. *Due to the de novo status of the variant in addition to the fitting clinical overlap to the rest of the cohort, this variant is deemed causative despite being classified as uncertain.

| Indi-vidual | Chr11: g.(hg38) | c. | p. | CADD-v1.6[2] | REVEL[3] | MutPred2[4] | VEST4[5] | BayesDel[6] | AA conservation (conserved up to) | gnomAD v4[7] |
|---|---|---|---|---|---|---|---|---|---|---|
| 1 | 67207624 | c.422A>G | p.(Tyr141Cys) | 32.0 | 0.836 | 0.522 | 0.858 | 0.389 | high (c. elegans) | 0 |
| 2 | 67217747 | c.704C>T | p.(Pro235Leu) | 34.0 | 0.880 | 0.594 | 0.821 | 0.426 | high (zebrafish) | 0 |
| 3 | 67219296 | c.850C>T | p.(His284Tyr) | 27.6 | 0.954 | 0.860 | 0.894 | 0.567 | high (c. elegans) | 0 |
| 4 | 67219402 | c.956G>A | p.(Arg319Gln) | 34.0 | 0.465 | 0.426 | 0.518 | 0.195 | high (c. elegans) | 0 |
| 5 | 67245196 | c.1571T>G | p.(Phe524Cys) | 21.0 | 0.040 | 0.334 | 0.286 | -0.099 | moderate (platypus) | 0 |
| 6 | 67245328 | c.1703G>A | p.(Arg568Gln) | 32.0 | 0.473 | 0.655 | 0.660 | 0.271 | high (c. elegans) | 0 |
| 7 | 67245397 | c.1772T>C | p.(Met591Thr) | 27.3 | 0.653 | 0.615 | 0.820 | 0.347 | high (zebrafish) | 0 |
| 8 | 67245421 | c.1796G>C | p.(Arg599Pro) | 33.0 | 0.531 | 0.611 | 0.800 | 0.314 | high (frog) | 0 |
| 9 | 67250357 | c.2327A>G | p.(Lys776Arg) | 23.8 | 0.096 | 0.298 | 0.272 | -0.081 | high (frog) | 0 |
| 10 | 67250358 | c.2328G>T | p.(Lys776Asn) | 23.5 | 0.109 | 0.313 | 0.381 | -0.164 | high (frog) | 0 |
| 11 | 67250461 | c.2431C>A | p.(His811Asn) | 22.1 | 0.104 | 0.479 | 0.298 | -0.234 | moderate (chicken) | 0 |

Table S2. *In silico* predition of missense variants in *KDM2A*.

MANE Select transcript NM_012308.3 was used. *In silico* scores were retrieved using dbNSFP[8]. Cutoffs for in silico scores were derived from Pejaver *et al.*[9] Red color signifies damaging prediction (at least PP3_SUP score in Pejaver *et al.* was reached); yellow color signifies neither PP3_SUP nor BP4_SUP score was reached; green color signifies benign prediction (at least BP4_SUP score in Pejaver *et al.* was reached). For further annotations on the missense variants, see Table S5 in separate excel file.

Table S3. Detailed clinical data of individuals with causative variants in *KDM2A*

See separate Excel file.

Table S4. Detailed clinical data of an individual with variant in *KDM2A* but insufficient evidence for causality

See separate Excel file.

Table S5. Annotation and *in silico* scores of all missense variants in *KDM2A*

See separate excel file. All annotations were retrieved using dbNSFP v4.5.[8]

Table S6. List of KDM genes

See separate Excel file.

Table S7. Reported variants in the literature in KDM genes

See separate Excel file.

Table S8. Methylation data of the *KDM2A*-related Episignature

See separate Excel file. Differential methylated regions between cases and control group. DMRs were filtered for an andjusted p-value of <0.01 and and minimal methylation difference of >10%.

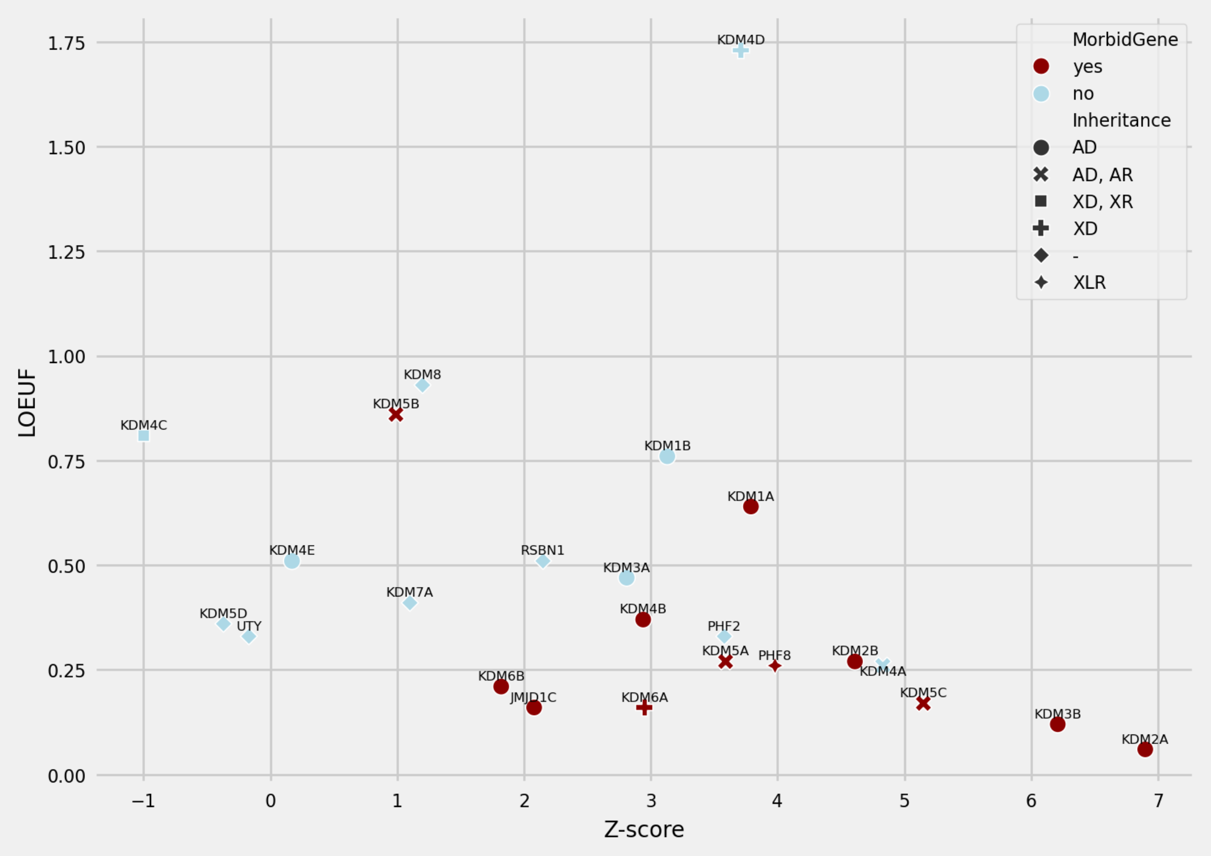


Figure S1. Constraint score landscape of KDM genes. Relationship between the LOEUF (predicted loss-of-function variants) and z-score (missense variants) among all KDM genes. KDM2A is the most constrained gene among this group with respect to both z-score and LOEUF. Constraint scores where preferably derived from gnomAD v4, but, if not available, also from gnomAD v2.1.1 (Table S5).[7]

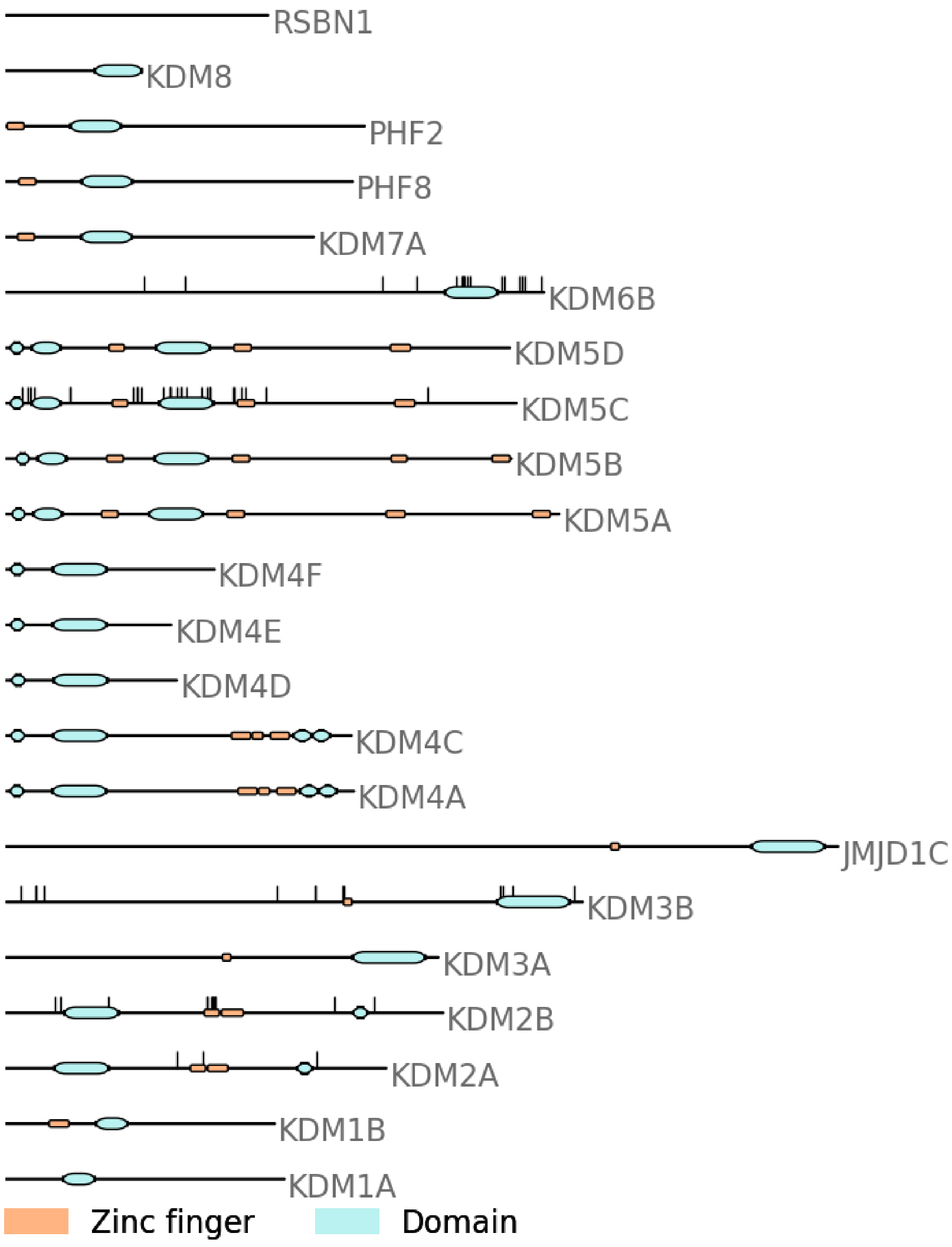


Figure S2. Overview of domain structure and reported variants in the literature in KDM genes. See Table S6 for a list of variants in KDM genes.

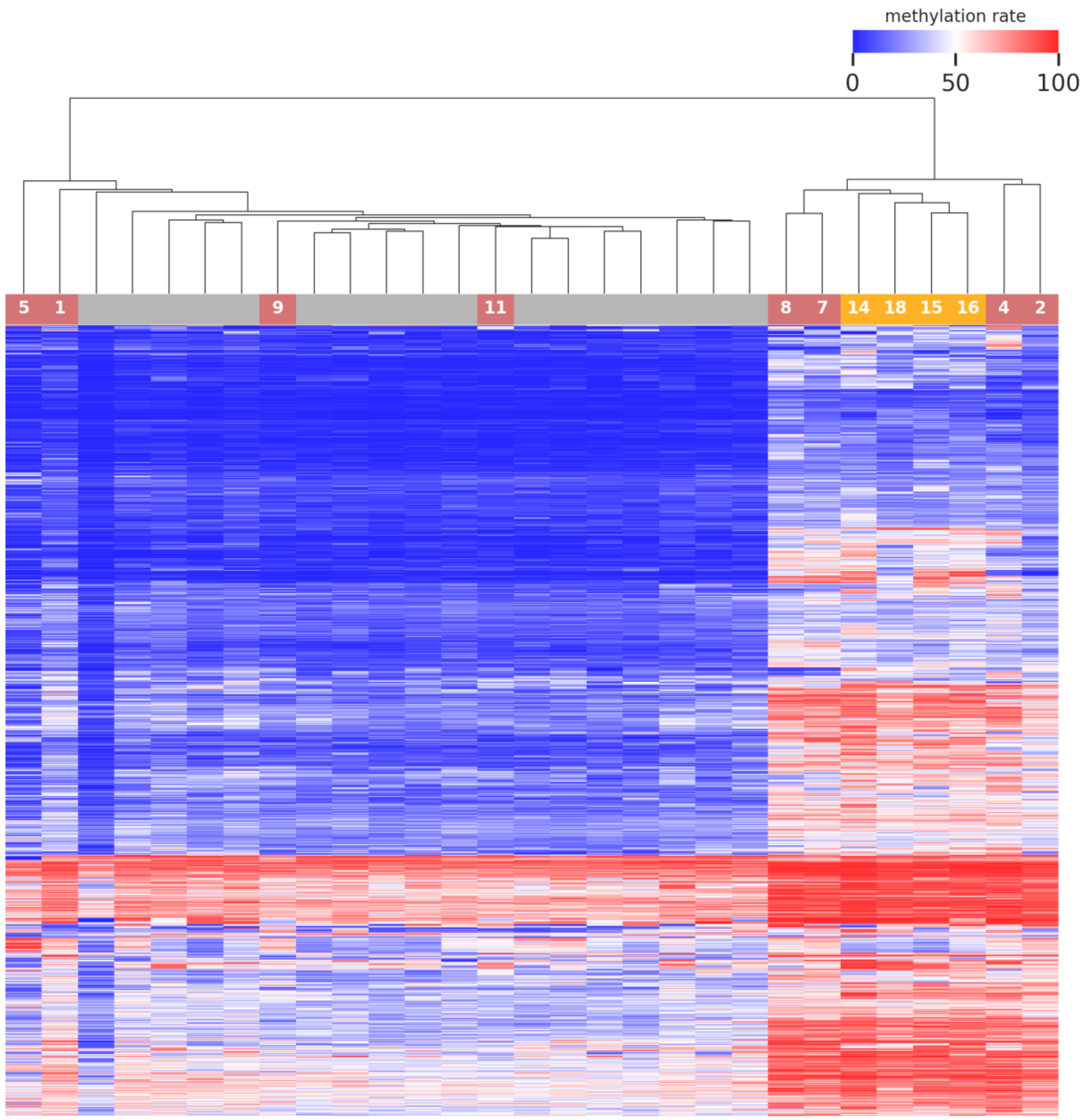


Figure S3. Methylation data of all variants. Heatmap displays hierarchical clustering of selected CpG sites of the episignature. Columns represent probes (grey: control probes; yellow: KDM2A pLoF variants; light red: KDM2A missense variants; number represents the individual in the cohort). Rows represent CpG sites. Color represents methylation ranging from dark blue (no methylation) to dark red (full methylation). A clear separation between control samples and a majority of samples of individuals with *de novo* variants in *KMD2A* is observed. Several individuals with *de novo* variants in *KDM2A* (individuals 1, 5, 9 and 11) cluster within the controls indicating no aberrant methylation profile.

#### References


1. Richards, S., Aziz, N., Bale, S., Bick, D., Das, S., Gastier-Foster, J., Grody, W.W., Hegde, M., Lyon, E., Spector, E., et al. (2015). Standards and guidelines for the interpretation of sequence variants: a joint consensus recommendation of the American College of Medical Genetics and Genomics and the Association for Molecular Pathology. Genetics in Medicine *17*, 405–424. 10.1038/gim.2015.30.

2. Rentzsch, P., Schubach, M., Shendure, J., and Kircher, M. (2021). CADD-Splice—improving genome-wide variant effect prediction using deep learning-derived splice scores. Genome Med *13*, 31. 10.1186/s13073-021-00835-9.

3. Ioannidis, N.M., Rothstein, J.H., Pejaver, V., Middha, S., McDonnell, S.K., Baheti, S., Musolf, A., Li, Q., Holzinger, E., Karyadi, D., et al. (2016). REVEL: An Ensemble Method for Predicting the Pathogenicity of Rare Missense Variants. The American Journal of Human Genetics *99*, 877–885. 10.1016/j.ajhg.2016.08.016.

4. Pejaver, V., Urresti, J., Lugo-Martinez, J., Pagel, K.A., Lin, G.N., Nam, H.-J., Mort, M., Cooper, D.N., Sebat, J., Iakoucheva, L.M., et al. (2020). Inferring the molecular and phenotypic impact of amino acid variants with MutPred2. Nat Commun *11*, 5918. 10.1038/s41467-020-19669-x.

5. Carter, H., Douville, C., Stenson, P.D., Cooper, D.N., and Karchin, R. (2013). Identifying Mendelian disease genes with the Variant Effect Scoring Tool. BMC Genomics *14*, S3. 10.1186/1471-2164-14-S3-S3.

6. Feng, B.-J. (2017). PERCH: A Unified Framework for Disease Gene Prioritization. Human Mutation *38*, 243–251. 10.1002/humu.23158.

7. Chen, S., Francioli, L.C., Goodrich, J.K., Collins, R.L., Kanai, M., Wang, Q., Alföldi, J., Watts, N.A., Vittal, C., Gauthier, L.D., et al. (2024). A genomic mutational constraint map using variation in 76,156 human genomes. Nature *625*, 92–100. 10.1038/s41586-023-06045-0.

8. Liu, X., Li, C., Mou, C., Dong, Y., and Tu, Y. (2020). dbNSFP v4: a comprehensive database of transcript-specific functional predictions and annotations for human nonsynonymous and splice-site SNVs. Genome Medicine *12*, 103. 10.1186/s13073-020-00803-9.

9. Pejaver, V., Byrne, A.B., Feng, B.-J., Pagel, K.A., Mooney, S.D., Karchin, R., O'Donnell-Luria, A., Harrison, S.M., Tavtigian, S.V., Greenblatt, M.S., et al. (2022). Calibration of computational tools for missense variant pathogenicity classification and ClinGen recommendations for PP3/BP4 criteria. The American Journal of Human Genetics *109*, 2163–2177. 10.1016/j.ajhg.2022.10.013.
